# Self-administered computerized cognitive training for cognitive deficits in individuals with metabolic syndrome: a randomized controlled trial

**DOI:** 10.64898/2026.06.22.26356249

**Authors:** Myrto Koutsonida, George Markozannes, Afroditi Kanellopoulou, Yiannis Tsiaras, Annita Varella, Vasiliki Zilidou, Evangelia Romanopoulou, Thomas Hyphantis, Evangelia Ntzani, Eleni Aretouli, Konstantinos K Tsilidis

## Abstract

**Background:** Metabolic syndrome (MetS) has been associated with cognitive decline. Considering its increasing prevalence worldwide, the goal of this study was to evaluate the feasibility and efficacy of a short-term, self-administered computerized cognitive training programme in individuals with metabolic syndrome and low cognitive performances.

**Methods:** Thirty six participants, aged 40-72 years (mean age: 57.8 years), were randomly assigned to the cognitive training or the passive control group. The cognitive training component of Long Lasting Memories (LLM) Care was used as an interactive software to enhance participants’ cognitive functions. Up to 24 sessions, each lasting 45 minutes, were self-administered at home twice per week for 3 months. Thorough cognitive assessments with were performed at baseline (randomization), at the end of intervention, and 12 months after baseline. The primary outcome was performance at nine neuropsychological tests, and the secondary outcome was a self-reported questionnaire assessing everyday functional abilities. Primary analyses were performed employing mixed-effect models using the intention-to-treat principle.

**Results:** Low adherence was observed in the study, as only 9 participants (50%) completed at least 8 sessions of the cognitive training programme (range 9-24 sessions, median 15 sessions). No statistically significant effect of the cognitive training programme on performance in neuropsychological tests or everyday functioning was found. At the end of the 3-month intervention programme, effect for visual memory enhancement in immediate (β = 1.58, 95% CI = -1.84 to 4.99, Cohen’s d = 0.39) and delayed recall (β = 2.17, 95% CI = -1.68 to 6.01, Cohen’s d = 0.45) was moderate in favour of the intervention group, and at 12-month follow-up, semantic verbal fluency gains for the intervention group were detected (β = 2.78, 95% CI = -0.92 to 6.49, Cohen’s d = 0.70), though with wide confidence intervals.

**Conclusions:** Despite some small effects observed in memory and verbal fluency, cognitive training did not yield statistically significant improvements. The observed low adherence and limited benefits on mild cognitive deficits in mostly middle-aged individuals with MetS are likely associated with the self-administered and short-term nature of the computerized intervention. This highlights the need for more intensive and clinician-delivered approaches to enhance engagement. Registry: ClinicalTrials.gov, TRN: NCT05658354, Registration date: 08 December 2022.

## Introduction

Metabolic syndrome (MetS) is a group of four cardiometabolic conditions, elevated blood pressure, glucose intolerance, dyslipidemia and abdominal obesity. Presence of MetS is considered a major risk factor for future cardiovascular diseases and has been associated with cognitive deficits in meta-analyses ^1,2^. Although there is still no consensus, it seems that individuals with MetS face impairments in attention ^3–5^, executive function ^6,7^ and memory ^8–10^. This association is particularly evident in middle-aged individuals ^11^, whereas a potential protective effect of MetS on cognitive function has been reported in older people Ill particularly for those aged above 75 years old ^12,13^ Ill, a finding that has been attributed to survival bias.

It is notable that even mild cognitive deficits can interfere with everyday activities ^14–16^, leading to reduced quality of life. For example, cognitive impairment was found to affect planning a driving route, speed and accuracy of financial transactions, telephone use and reading concentration. Consequently, the cognitive deficits associated with MetS, even when subtle, may have meaningful repercussions for daily functioning and overall well-being.

Moreover, growing evidence indicates that cognitive dysfunction increases the likelihood of progression to mild cognitive impairment (MCI) and, eventually, dementia in older adults ^17–19^ and also in individuals with MetS ^20–22^. As in some persons, MCI is potentially reversible when detected and treated at an early stage, identifying and addressing cognitive decline at pre-clinical stages has become an important public health priority.

Over the past years, cognitive training has gained significant attention as a potential intervention to maintain or improve cognitive function. Cognitive training refers to a programme of standardized exercises aimed to train specific cognitive functions, including attention, executive functions and memory. The programme can be unimodal, focusing in one distinct cognitive function (e.g memory), or multimodal, focusing on more cognitive functions, and can be delivered paper-and-pencil or computer-based ^23^.

Despite that cognitive training has been shown to be effective in healthy older adults ^24–26^, individuals with psychiatric disorders ^27–29^, neurocognitive disorders ^30–33^ and other neurological conditions ^34–36^, little is known about the potential of this intervention to address subtle cognitive deficits linked with the presence of MetS. As prevalence of MetS is growing worldwide ^37^, it is critical to explore the possibility of enhancing cognitive abilities in individuals with MetS using cognitive training.

The aim of the present study was to investigate the feasibility and efficacy of a 3-month self-administered computerized cognitive training programme in individuals with MetS and subtle cognitive deficits. Outcomes were compared between a training and a passive control group upon completion of the 3-month intervention and 12 months after the baseline examination. It was hypothesized that the intervention would be associated with improved cognitive performance at post-intervention and that these improvements would be maintained at the 12-month follow-up. A secondary aim of the current study was to evaluate the impact of the cognitive training programme on self-reported quality of life.

## Methods

### Study participants

Participants were primarily recruited from the Epirus Health Study (EHS) cohort (https://ehs.med.uoi.gr/), an innovative prospective study designed to investigate the aetiology of complex multifactorial chronic diseases in Greece and to improve the overall health state of the Greek population. It consists of permanent residents of the northwest region of Epirus in Greece, aged 20–80 years. The EHS was initiated in June 2019, and the recruitment was completed in October 2023. Until then, 2,540 community adults were recruited.

Details of EHS have been published elsewhere ^38,39^. Briefly, the EHS collected information on socio-demographic characteristics, lifestyle data, anthropometric, biochemical, clinical and cognitive measurements. At recruitment, participants underwent an interview and a clinical examination by two trained medical professionals. Basic demographic characteristics (i.e., age, sex, place of birth, marital status, level of education, current employment status and income), personal and family medical history, and lifestyle factors (i.e., physical activity, smoking habits, alcohol consumption) were assessed using a standard questionnaire. Weight, standing height and waist circumference were measured using SECA equipment. Systolic and diastolic blood pressures were measured using the MicroLife A6 PC-AFIB PC monitor. Cognitive functions were assessed using the paper-based Greek versions of the Trail Making Test ^40^, Verbal Fluency test ^41^ and Logical Memory test ^42^.

Given the absence of randomized controlled trials evaluating computerized cognitive training in individuals with coexisting mild cognitive impairment and metabolic syndrome, a total sample of 50 participants (25 per group) was selected in accordance with recommendations for pilot randomized controlled trials^43^.

Due to the limited number of EHS participants meeting the study’s inclusion criteria, additional participants were recruited through self-referrals from participants’ social networks. All recruited individuals underwent a screening process to confirm eligibility prior to enrollment.

Eligibility criteria were: (1) Presence of MetS according to International Diabetes Federation (IDF) criteria ^44^ or revised National Cholesterol Education Program-Adult Treatment Panel III (NCEP-ATP III) criteria ^45^, and (2) Presence of MCI, determined psychometrically according to the Peterson criteria ^46^. Participants were excluded if they self-reported any serious neurological or psychiatric condition.

According to the revised NCEP-ATP III criteria, MetS is diagnosed when at least three of the following conditions are met: (a) waist circumference ≥102 cm in men or ≥88 cm in women, (b) triglycerides ≥150 mg/dL or lipid lowering drug treatment, (c) high-density lipoprotein (HDL) cholesterol <40 mg/dL in men or <50 mg/dL in women or lipid lowering drug treatment, (d) systolic blood pressure ≥130 mm Hg or diastolic blood pressure ≥85 mm Hg or anti-hypertensive drug treatment, (e) fasting glucose ≥100 mg/dL or anti-diabetic drug treatment. The IDF criteria require the presence of central obesity, defined by ethnic-specific waist circumference (Europeans ≥94 cm in men or ≥80 in women), or a body mass index (BMI) over 30 kg/m^2^, plus any two of the following: (a) triglycerides ≥150 mg/dL or drug treatment, (b) HDL cholesterol <40 mg/dL in men or <50 mg/dL in women or drug treatment, (c) systolic blood pressure ≥130 mm Hg or diastolic blood pressure ≥85 mm Hg or anti-hypertensive drug treatment, (d) fasting glucose ≥100 mg/dL or previously diagnosed type 2 diabetes.

MCI was defined as performance at least 1.5 standard deviation (SD) below published age- and education-adjusted normative data for the Greek population on one or more of the following previously administered neuropsychological tests: Trail Making Test, Verbal Fluency test or Logical Memory test.

Eligible participants were contacted through telephone and invited to participate by the study’s neuropsychologist. The recruitment for the current clinical trial was accomplished from December 2022 to March 2024. All participants provided written informed consent prior to participation in the study.

The study’s protocol was approved by the Research Ethics Committee of the University of Ioannina (27819/25-05-2022), and was conducted in accordance with the Declaration of Helsinki. The clinical trial’s protocol was registered at clinicaltrials.gov (NCT05658354).

### Study procedures

In the initial baseline assessment (T0), participants completed a questionnaire that verified and updated previous collected information on socio-demographic and lifestyle characteristics (age, education, income, physical activity, smoking, alcohol consumption, height, weight, waist circumference, and current medication).

After baseline assessment, participants were randomly assigned to one of the two study arms, an intervention group that received the cognitive training programme or a passive control group. Randomization was carried out sing a computer-generated sequence in blocks of four with a 1:1 ratio. Blinding of test administrator was not feasible due to practical issues whereas participants were blinded to group allocation. All participants, regardless of treatment group, received the World Health Organization (WHO) guidelines on risk reduction of cognitive decline and dementia ^47^.

Then, a comprehensive battery of paper-and-pencil and computerized neuropsychological tests, as well as a questionnaire on everyday functioning, were administered by the study’s neuropsychologist. Subsequent neuropsychological assessments occurred at the end of the 3-month cognitive training period (T1), and 12 months after the baseline assessment (T2). A questionnaire on daily functioning was administered again at the final assessment (T2).

All assessments were performed at the University of Ioannina.

### Cognitive training programme

Cognitive training was delivered using the web-based cognitive training component of Long Lasting Memories (LLM) Care. This is a Greek version of the BrainHQ software (BrainHQ, Posit Sciences, https://www.brainhq.com), adapted in the Greek language and cultural context by Bamidis et al. ^48^. LLM Care is considered a non-pharmacological intervention for mitigating cognitive decline, providing structured training to individuals at increased risk. It consists of 29 tasks targeting attention, brain speed, memory, people skills, navigation and intelligence. Task difficulty is continuously adjusted to each participant’s performance, enabling a personalized training experience.

Based on the specific cognitive deficits of participants with MetS ^39^, 10 tasks were selected to train episodic memory (Face Facts, In the Know), memory and navigation (Mental Map, True North), working memory (Auditory Ace, Card Shark), and attention and processing speed (Divided Attention, Optic Flow, Mixed Signals, Sound Sweeps). Further details for each task are provided in the supplementary material (Supplementary Table S1).

Participants assigned to the intervention group were instructed to complete a total of 24 sessions, two sessions per week, for 3 months. Each session was planned to last 45 minutes. Participants were trained for the use of the platform by the study’s neuropsychologist and received a training manual with detailed instructions for each task. The cognitive training programme was implemented at home according to the pre-specified schedule. Participation was tracked remotely by the study’s neuropsychologist via the programme’s automated logs.

In cases of non-adherence, participants received a weekly reminder email encouraging them to follow the programme. If non-adherence persisted, the study’s neuropsychologist contacted the participants by phone to provide additional support and promote adherence. Furthermore, participants in the intervention group received monthly telephone calls to address and resolve any potential technical issues related to the programme.

### Study Outcome Measures

At T0, T1, and T2, both paper-and-pencil and computerized neuropsychological tests were administered to all participants to evaluate the efficacy of the cognitive training programme. The neuropsychological battery was comprised of standardized neuropsychological tests using original versions along with equivalent, alternate versions to control for practice effects. Some tests originally specified in the protocol (e.g., the Clock Drawing Test) were substituted with more comprehensive measures (e.g., NIH-EXAMINER battery) assessing the same cognitive domains with greater sensitivity. These modifications were implemented prior to data collection to improve psychometric properties and did not affect the predefined cognitive domains assessed.

The paper-and-pencil neuropsychological tests included the Greek adaptations of the Montreal Cognitive Assessment (MoCA) ^49^ as a screening tool for general cognitive status, the Hopkins Verbal Learning Test-Revised (HVLT-R) ^50^ as a test of verbal memory, the Taylor Complex Figure ^51^ and the Georgia Complex Figure 1 and 2 tests^52^ as measurments of visuo-construction abilities and visual memory, the Digit Span forward and backward condition ^42^ as a test of attention and working memory, and the semantic Verbal Fluency ^41^ (fruits, jobs, objects) as a test of executive function that also relies on language.

**Figure 1.**
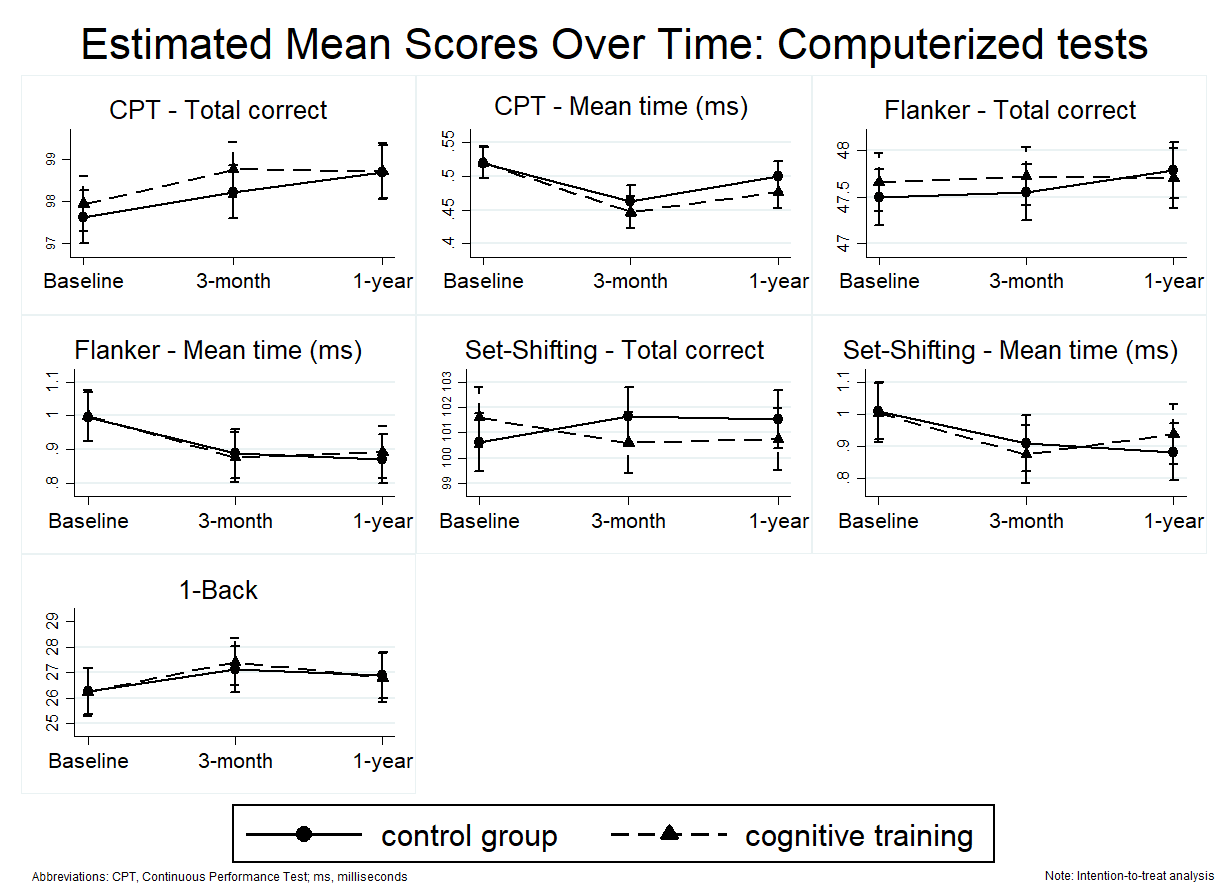
Flowchart of participants

**Figure 2.**
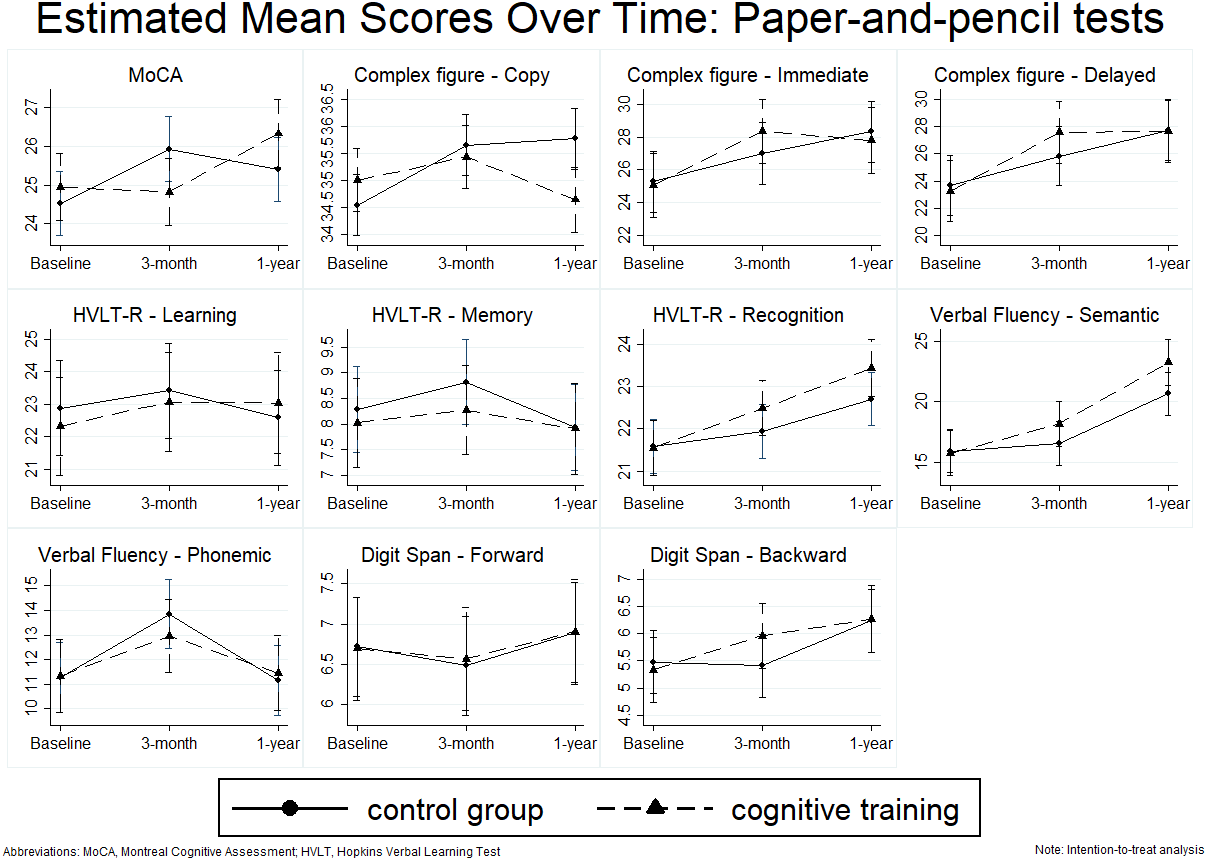
Estimated means of paper-and-pencil cognitive scores over time by intervention group according to intention-to-treat principle. Estimated marginal means for the cognitive training and control group at baseline, 3 months, and 1 year derived from linear mixed-effects models. Error bars represent 95% confidence intervals.

Computerized neuropsychological tests included 4 tasks from NIH-EXAMINER battery ^53^, a validated battery developed by the National Institutes of Health (NIH) to assess executive functions. Namely, the 4 tasks administered were the Flanker task and the Continuous Performance Test (CPT) as tests of inhibition, the Set-Shifting task as test of flexibility, and the N-Back task as a test of working memory.

In all the above-mentioned paper-and-pencil and computerized neuropsychological tests, the sum of correct responses was calculated, with higher scores indicating better performance. In three of the computerized neuropsychological tests (Flanker, CPT and Set-Shifting), we also calculated the mean reaction time in milliseconds (ms). Lower reaction time indicated better performance.

In addition, the revised Everyday Cognition scale (ECog-II) ^54^ was administered at T0 and T2, to assess functional abilities as a proxy of quality of life. The ECog-II is a 41-item measure of everyday functioning that evaluates memory, language, visual, spatial and perceptual abilities, planning, organization, and divided attention. Each item is rated on a 4-point Likert scale: 1 = better or no change; 2 = occasionally worse/questionable; 3 = consistently a little worse; 4 = consistently much worse, with an additional option available for cases where the respondent is unable to provide an answer (“Don’t know/Not applicable”). Total scores were used in the analyses, with higher scores indicating greater impairment in everyday functioning. This instrument was introduced prior data collection in place of the SF-36 Health Survey originally specified in the protocol, to provide a domain-specific assessment of everyday cognitive functioning more closely aligned with the intervention targets.

### Statistical analysis

Baseline socio-demographic characteristics and cognitive scores of the study participants were summarized using means and SD for continuous variables, and percentages for categorical variables. Independent sample t-tests or Fisher’s exact tests were applied to compare the baseline characteristics between the two groups.

Separate mixed-effects models were fitted for each of the nine neuropsychological tests to assess differences in scores between the two groups across the three timepoints. Models were fitted with a random intercept per participant and fixed effects of intervention group, timepoint and timepoint-by-intervention group interaction. Models were adjusted for baseline cognitive score (continuous), age (continuous), sex, education (primary and secondary school, high school, higher education), and recreational physical activity [measured in Metabolic Equivalents of Task (MET) per hour/week] (continuous). Linear mixed-effects models were used instead of the ANCOVA approach specified in the original protocol, as they allow the inclusion of all available repeated measurements and provide a more flexible approach for handling missing data under the missing-at-random assumption.

Between-group effect sizes (Cohen’s d) were calculated from model-adjusted means and pooled residual standard deviations obtained from the mixed-effects models. Effect sizes of 0.2, 0.5, and 0.8 were interpreted as small, medium, and large, respectively.

Pairwise comparisons were performed using Wald test to compare cognitive performance within groups at each follow-up timepoint.

Mixed-effects models were also used to assess differences between the two groups in self-reported everyday functional abilities, as measured by the ECog-II questionnaire, from baseline (T0) to the 12-month follow-up (T2). Models were fitted with a random intercept per participant, fixed effects of intervention group, timepoint and timepoint-by-intervention group interaction, and were adjusted for age, sex, education and recreational physical activity.

In the primary analyses, data were analyzed according to the intention-to-treat principle and all participants were included in the group they were originally assigned to, regardless of adherence to the treatment protocol. In the secondary analyses, we followed an as-treated principle, including participants in the intervention group only if they had completed at least 8 sessions of the cognitive training programme at the training site, regardless of the timing of these sessions.

All statistical analyses were undertaken using STATA (version 14; StataCorp, College Station, TX, USA). After applying the Bonferroni correction for the ten individual tests conducted (nine neuropsychological tests and one functional abilities questionnaire), the adjusted significance threshold was set at 0.005.

## Results

### Participants’ characteristics and intervention participation

Of the 92 eligible participants, 36 consented to participate and were randomized, with 18 participants initially allocated in the intervention group and 18 in the control group (Figure 1).

Supplementary Table S2. presents the socio-demographic characteristics of enrolled and non-enrolled participants, who had similar characteristics except for two components of MetS, namely higher hypertension and hyperglycemia proportions in the enrolled participants. Of the enrolled participants, 3 were lost to follow-up after baseline assessment and one was lost during the second follow-up period (T2). The reasons for these drop-outs were time commitment (1 intervention, 1 control), lack of motivation (1 intervention), or not provided (1 intervention). Participants who dropped out did not differ significantly from those that completed the study in terms of age, sex, education, MetS components, smoking, alcohol consumption, income and cognitive function (Supplementary Table S3).

Table 1 presents the baseline socio-demographic characteristics and the baseline cognitive scores of the study participants overall and by group, according to the intention-to-treat principle. The mean age of study participants was 57.8 years (SD=8.0). Women (57.6%) and individuals of higher education (60.6%) and of higher income (69.7%) predominated in the sample. The most frequent MetS component was dyslipidemia (87.9%), and the least frequent was hyperglycemia (33.3%). The total sample consisted of 16 participants in the intervention group and 17 participants in the control group. Groups did not differ significantly on baseline socio-demographic and cognitive variables with the exception of the Set-Shifting task, where the intervention group outperformed the control group (mean difference = 2.19, p = 0.034). Similar results were obtained when baseline characteristics were compared between the groups according to as-treated principle (Supplementary Table S4).

**Table 1.**
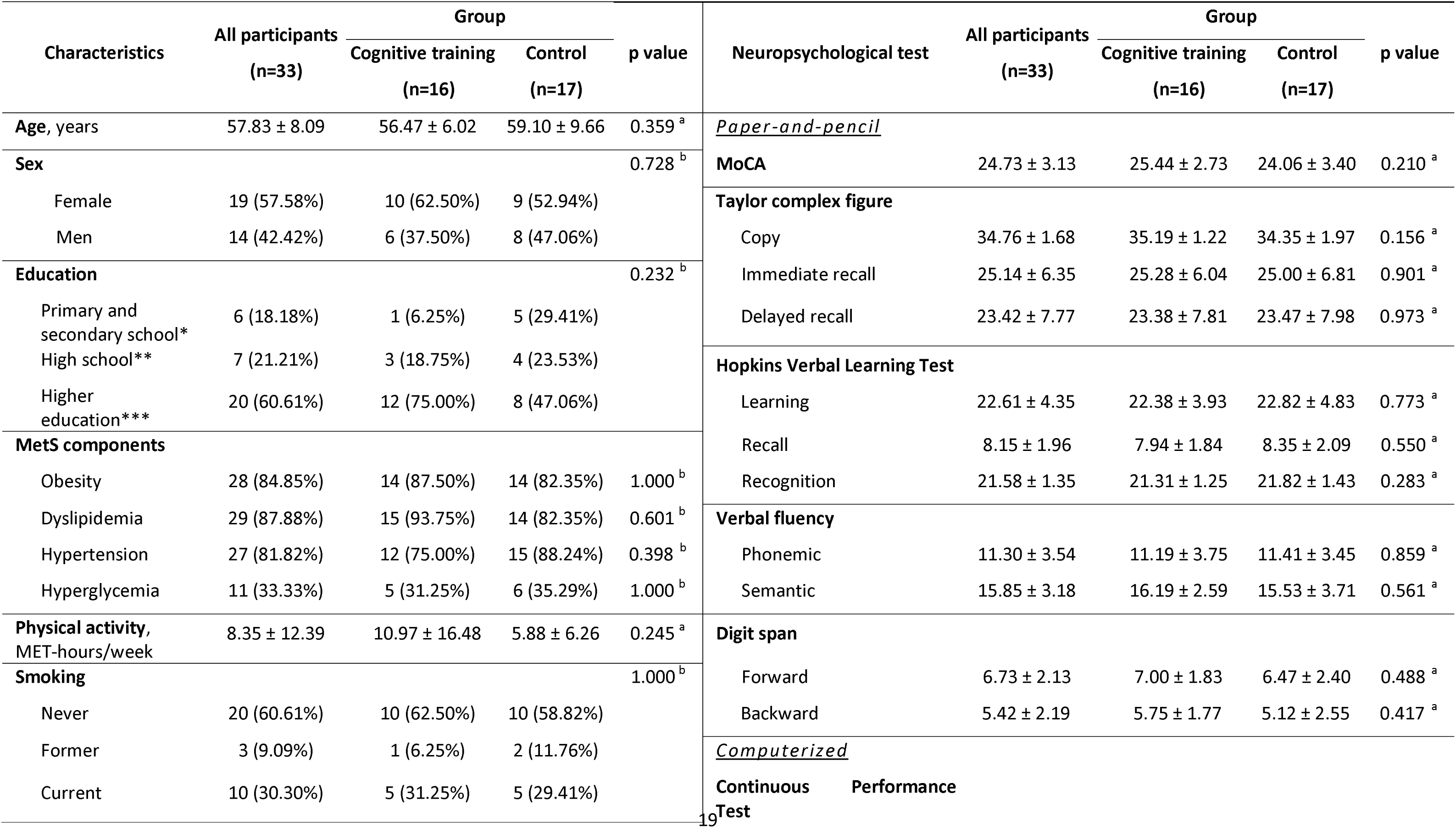
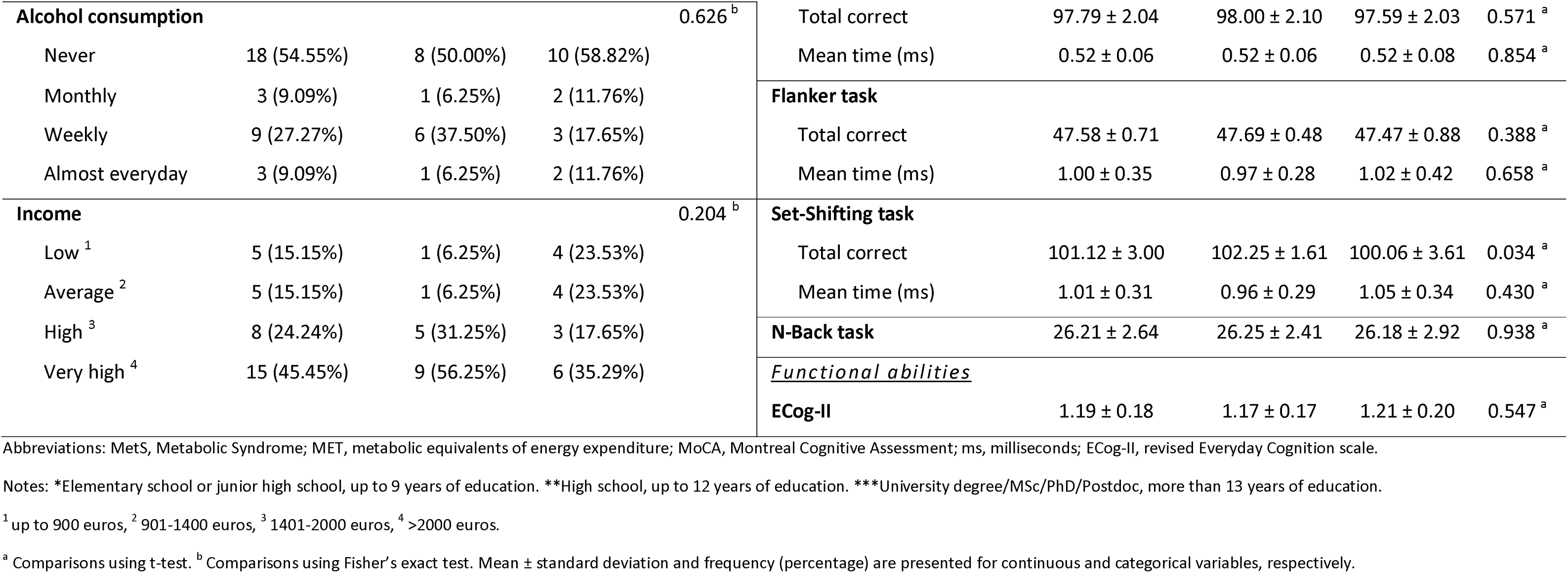
Baseline socio-demographic characteristics and cognitive scores of study participants according to intention-to-treat principle.

In total, 9 of the initially allocated participants (50%) completed at least 8 sessions and were therefore classified as adherent to the intervention programme (range 9-24 sessions, median 15 sessions). Non-adherent participants, who did not complete 8 sessions of 45-minute cognitive training, were equivalent to adherent participants in baseline socio-demographic and cognitive variables (Supplementary Table S5). However, older participants tended to complete more cognitive training sessions (r = 0.46, p = 0.045) (Supplementary Figure S1).

### Cognitive training

Supplementary Table S6. reports the means and standard deviations for cognitive test scores at pre- and post-intervention timepoints by treatment group according to the intention-to-treat principle. Corresponding intention-to-treat results are shown in Table 2 and Figures 2 and 3. Analyses using mixed-effects models that take in consideration all time-points of outcome assessment showed no statistically significant effect of timepoint-by-intervention group interaction, indicating that difference in scores is roughly the same for both the intervention and the control group at the two follow-up timepoints. The interaction found in the copy condition of the complex figure was not significant after Bonferroni correction.

**Figure 3.**
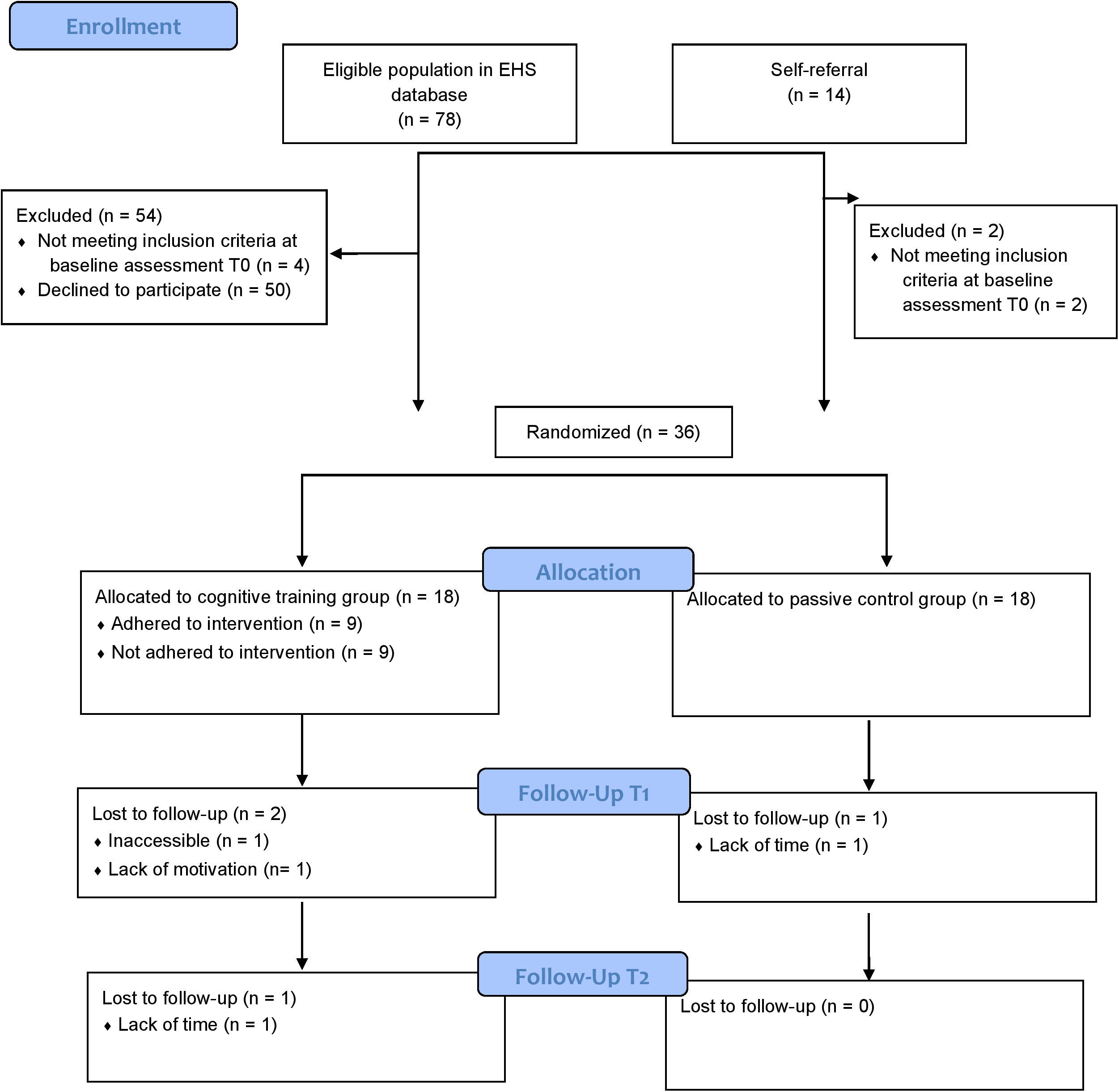
Estimated means of computerized cognitive scores over time by intervention group according to intention-to-treat principle. Estimated marginal means for the cognitive training and control group at baseline, 3 months, and 1 year derived from linear mixed-effects models. Error bars represent 95% confidence intervals.

**Table 2.** Mixed models results for cognitive scores according to intention-to-treat principle.

| Neuropsychological test | Beta | 95% CI | Effect size, *d* | Neuropsychological test | Beta | 95% CI | Effect size, *d* |
| --- | --- | --- | --- | --- | --- | --- | --- |
| <u>Paper-and-pencil</u> |  |  |  | <u>Computerized</u> |  |  |  |
| **MoCA** |  |  |  | **CPT - Total correct** |  |  |  |
| 3-month | -1.54 | -3.23, 0.16 | 0.64 | 3-month | 0.22 | -1.05, 1.50 | 0.41 |
| 1-year | 0.50 | -1.21, 2.21 | 0.54 | 1-year | -0.30 | -1.58, 0.99 | 0.01 |
| **Complex figure - Copy** |  |  |  | **CPT - Mean time (ms)** |  |  |  |
| 3-month | -0.68 | -1.83, 0.47 | 0.19 | 3-month | -0.02 | -0.06, 0.03 | 0.34 |
| 1-year | -1.59* | -2.75, -0.43 | 0.97 | 1-year | -0.03 | -0.07, 0.02 | 0.51 |
| **Complex figure - Immediate** |  |  |  | **Flanker task - Total correct** |  |  |  |
| 3-month | 1.58 | -1.84, 4.99 | 0.39 | 3-month | 0.00 | -0.61, 0.62 | 0.27 |
| 1-year | -0.30 | -3.75, 3.14 | 0.15 | 1-year | -0.25 | -0.87, 0.37 | 0.14 |
| **Complex figure - Delayed** |  |  |  | **Flanker task - Mean time (ms)** |  |  |  |
| 3-month | 2.17 | -1.68, 6.01 | 0.45 | 3-month | -0.01 | -0.16, 0.13 | 0.00 |
| 1-year | 0.36 | -3.52, 4.25 | 0.01 | 1-year | 0.02 | -0.13, 0.16 | 0.00 |
| **HVLT - Learning** |  |  |  | **Set-Shifting task - Total correct** |  |  |  |
| 3-month | 0.22 | -2.76, 3.20 | 0.12 | 3-month | -0.75 | -2.65, 1.15 | 0.21 |
| 1-year | 1.01 | -1.99, 4.02 | 0.15 | 1-year | -0.77 | -2.68, 1.15 | 0.21 |
| **HVLT - Recall** |  |  |  | **Set-Shifting task - Mean time (ms)** |  |  |  |
| 3-month | -0.28 | -1.79, 1.23 | 0.35 | 3-month | 0.02 | -0.15, 0.17 | 0.08 |
| 1-year | 0.24 | -1.28, 1.77 | 0.01 | 1-year | 0.10 | -0.06, 0.27 | 0.63 |
| **HVLT - Recognition** |  |  |  | **N-Back task** |  |  |  |
| 3-month | 0.59 | -0.68, 1.85 | 0.43 | 3-month | 0.31 | -1.54, 2.15 | 0.16 |
| 1-year | 0.77 | -0.50, 2.04 | 0.58 | 1-year | -0.09 | -1.95, 1.77 | 0.06 |
| **Verbal fluency - Phonemic** |  |  |  |  |  |  |  |
| 3-month | -0.96 | -3.58, 1.65 | 0.34 |  |  |  |  |
| 1-year | 0.24 | -2.40, 2.88 | 0.12 |  |  |  |  |
| **Verbal fluency - Semantic** |  |  |  |  |  |  |  |
| 3-month | 1.79 | -1.88, 5.46 | 0.44 |  |  |  |  |
| 1-year | 2.78 | -0.92, 6.49 | 0.70 |  |  |  |  |
| **Digit span - Forward** |  |  |  |  |  |  |  |
| 3-month | 0.11 | -1.00, 1.22 | 0.08 |  |  |  |  |
| 1-year | 0.03 | -1.09, 1.16 | 0.01 |  |  |  |  |
| **Digit span - Backward** |  |  |  |  |  |  |  |
| 3-month | 0.68 | -0.42, 1.79 | 0.49 |  |  |  |  |
| 1-year | 0.16 | -0.95, 1.28 | 0.03 |  |  |  |  |

Abbreviations: CI, confidence interval; MoCA, Montreal Cognitive Assessment; HVLT, Hopkins Verbal Learning Test; CPT, Continuous Performance Test; ms, milliseconds.
Notes: Models adjusted for baseline score, age, sex, education and recreational physical activity. p values represent statistical significance of the group x timepoint interaction term.
\* p <0.05, \*\*p < 0.003

In general, small to moderate improvements across several domains were noted for the intervention group both upon completion of the 3-month intervention and at 12-month follow-up, but with wide confidence intervals. Large effect estimates favouring the intervention group were observed at the 3-month period in the memory conditions of the complex figures test (immediate recall: β = 1.58, 95% CI = -1.84 to 4.99, d = 0.39; delayed recall: β = 2.17, 95% CI = -1.68 to 6.01, d = 0.45), and at both follow-up periods in the semantic condition of the Verbal Fluency test (3-month: β = 1.79, 95% CI = -1.88 to 5.46, d = 0.44; 12-month: β = 2.78, 95% CI = -0.92 to 6.49, d = 0.70). In the copy condition of the complex figures (12-month: β = -1.59, 95% CI = -2.75 to -0.43, d = 0.97) and in the MoCA test (3-month: β = -1.54, 95% CI = -3.23 to 0.16, d = 0.64), large effect estimates favouring the control group were seen, but they may reflect transient fluctuations as they were not consistent with the rest of the findings. Performance at the computerized neuropsychological tests showed little change between the two follow-up periods.

Because of the low adherence rate, mixed-effects model analyses were repeated according to the as-treated principle, including only participants who completed at least 8 sessions of the cognitive training programme in the intervention group. Results remained fairly unchanged (Supplementary Table S7, Supplementary Figures S2-S3). Notably, when only adherent participants were considered, the largest effect estimates favouring the intervention group emerged mostly in memory tests. More specifically, at both follow-up timepoints in immediate recall (3-month: β = 3.02, 95% CI = -0.78 to 6.82, d = 1.03; 12-month: β = 1.39, 95% CI = -2.51 to 5.29, d = 0.55) and delayed recall of the complex figures test (3-month: β = 3.85, 95% CI = -0.39 to 8.10, d = 1.07; 12-month: β = 3.02, 95% CI = -1.34 to 7.38, d = 0.86), and at 12-month follow-up period in the HVLT-R (learning: β = 1.77, 95% CI = -1.60 to 5.14, d = 0.73; memory: β = 1.66, 95% CI = -0.03 to 3.36, d = 0.89; recognition: β = 0.93, 95% CI = -0.52 to 2.37, d = 0.84).

Supplementary Figure S4 depicts the associations between age, cognitive performance at the 3-month follow-up, and total training hours. There appears to be a positive trend between training duration and test scores, particularly in memory tests.

When we performed analyses within study groups, both groups showed improvements on selective memory tests, semantic verbal fluency and processing speed measurements (Table 3). Regarding the paper-and-pencil neuropsychological tests, improvements were demonstrated on the delayed recall of the complex figure at 3-month for the intervention group (MD = 4.31, p = 0.027) and at 12-month for the control group (MD = 4.03, p = 0.039), and on the semantic condition of the Verbal Fluency test at 12-month follow-up for both groups (cognitive training: MD = 7.60, p < 0.001; control: MD = 4.77, p = 0.003). Moreover, significant gains were found at the 12-month follow-up on the recognition condition of the HVLT-R in the intervention group (MD = 1.93, p = 0.001). Regarding the computerized neuropsychological tests, significant reductions in reaction time were seen for both groups mainly at 3-month in CPT (cognitive training: MD = -0.07, p < 0.001; control: MD = -0.06, p = 0.005), but these improvements were not maintained at 12-month. These results exhibited little variations when analyses were repeated according to as-treated principle (Supplementary Table S8).

**Table 3.** Mean change in cognitive scores at 3-month and 1-year follow-up within groups according to intention-to-treat principle.

| Neuropsychological test | Cognitive training (n=16) |  | Control (n=17) |  | Neuropsychological test | Cognitive training (n=16) |  | Control (n=17) |  |
| --- | --- | --- | --- | --- | --- | --- | --- | --- | --- |
|  | MD (SD) | p value | MD (SD) | p value |  | MD (SD) | p value | MD (SD) | p value |
| <u>Paper-and-pencil</u> |  |  |  |  | <u>Computerized</u> |  |  |  |  |
| **MoCA** |  |  |  |  | **CPT - Total correct** |  |  |  |  |
| 3-month | -0.13 (2.75) | 1.000 | 1.41 (3.10) | 0.253 | 3-month | 0.81 (1.11) | 1.000 | 0.59 (2.74) | 1.000 |
| 1-year | 1.33 (2.32) | 0.385 | 0.88 (2.12) | 1.000 | 1-year | 0.80 (1.86) | 1.000 | 1.06 (2.08) | 0.251 |
| **Complex figure - Copy** |  |  |  |  | **CPT - Mean time (ms)** |  |  |  |  |
| 3-month | 0.44 (1.63) | 1.000 | 1.12 (1.76) | 0.079 | 3-month | -0.07 (0.06) | <0.001 | -0.06 (0.07) | 0.005 |
| 1-year | -0.40 (2.20) | 0.880 | 1.24 (1.44) | 0.030 | 1-year | -0.04 (0.04) | 0.132 | -0.02 (0.06) | 1.000 |
| **Complex figure - Immediate** |  |  |  |  | **Flanker task - Total correct** |  |  |  |  |
| 3-month | 3.28 (4.24) | 0.434 | 1.71 (4.29) | 1.000 | 3-month | 0.06 (0.77) | 1.000 | 0.06 (1.25) | 1.000 |
| 1-year | 2.70 (5.79) | 1.000 | 3.03 (4.96) | 0.161 | 1-year | 0.07 (0.80) | 1.000 | 0.29 (0.85) | 1.000 |
| **Complex figure - Delayed** |  |  |  |  | **Flanker task - Mean time (ms)** |  |  |  |  |
| 3-month | 4.31 (5.57) | 0.027 | 2.15 (6.23) | 1.000 | 3-month | -0.12 (0.10) | 0.244 | -0.11 (0.27) | 0.431 |
| 1-year | 4.23 (5.82) | 1.000 | 4.03 (5.70) | 0.039 | 1-year | -0.11 (0.16) | 0.564 | -0.12 (0.28) | 0.185 |
| **HVLT-R - Learning** |  |  |  |  | **Set-Shifting task - Total correct** |  |  |  |  |
| 3-month | 0.75 (3.30) | 1.000 | 0.53 (5.76) | 1.000 | 3-month | -0.75 (1.92) | 1.000 | 0.00 (2.83) | 1.000 |
| 1-year | 0.60 (3.80) | 1.000 | -0.29 (4.04) | 1.000 | 1-year | -0.67 (2.16) | 1.000 | 0.06 (2.99) | 1.000 |
| **HVLT-R - Recall** |  |  |  |  | **Set-Shifting task - Mean time (ms)** |  |  |  |  |
| 3-month | 0.25 (1.77) | 1.000 | 0.53 (2.27) | 1.000 | 3-month | -0.17 (0.14) | 0.036 | -0.19 (0.28) | 0.011 |
| 1-year | -0.20 (2.24) | 1.000 | -0.35 (2.32) | 1.000 | 1-year | -0.08 (0.15) | 1.000 | -0.19 (0.29) | 0.008 |
| **HVLT-R - Recognition** |  |  |  |  | **N-Back task** |  |  |  |  |
| 3-month | 0.94 (1.48) | 0.577 | 0.35 (1.73) | 1.000 | 3-month | 1.19 (2.86) | 1.000 | 0.88 (3.22) | 1.000 |
| 1-year | 1.93 (1.62) | 0.001 | 1.12 (1.53) | 0.164 | 1-year | 0.27 (1.87) | 1.000 | 0.65 (3.33) | 1.000 |
| **Verbal fluency - Phonemic** |  |  |  |  |  |  |  |  |  |
| 3-month | 1.63 (4.79) | 1.000 | 2.59 (2.98) | 0.068 |  |  |  |  |  |
| 1-year | 0.20 (3.21) | 1.000 | -0.12 (3.55) | 1.000 |  |  |  |  |  |
| **Verbal fluency - Semantic** |  |  |  |  |  |  |  |  |  |
| 3-month | 2.44 (4.86) | 0.964 | 0.65 (4.03) | 1.000 |  |  |  |  |  |
| 1-year | 7.60 (5.55) | <0.001 | 4.77 (4.87) | 0.003 |  |  |  |  |  |
| **Digit span - Forward** |  |  |  |  |  |  |  |  |  |
| 3-month | -0.13 (1.36) | 1.000 | -0.24 (1.35) | 1.000 |  |  |  |  |  |
| 1-year | 0.20 (1.74) | 1.000 | 0.18 (1.85) | 1.000 |  |  |  |  |  |
| **Digit span - Backward** |  |  |  |  |  |  |  |  |  |
| 3-month | 0.63 (1.67) | 1.000 | -0.06 (1.68) | 1.000 |  |  |  |  |  |
| 1-year | 0.93 (1.79) | 0.320 | 0.77 (0.97) | 0.698 |  |  |  |  |  |

Abbreviations: MD, mean difference; SD, standard deviation; MoCA, Montreal Cognitive Assessment; HVLT, Hopkins Verbal Learning Test; CPT, Continuous Performance Test; ms, milliseconds.
Notes: Models adjusted for baseline score, age, sex, education and recreational physical activity. p values represent statistical significance derived from Wald test.

Mixed-effects models were conducted to examine differences in the self-reported everyday functioning between groups. No statistically significant difference was found (Table 4), neither according to the intention-to-treat principle (β = 0.02, p = 0.791) nor according to the as-treated principle (β = 0.02, p = 0.800).

**Table 4.** Mixed models results for scores of everyday functioning between groups.

| ECog- II score | Beta | 95% CI | p value |
| --- | --- | --- | --- |
| <u>*Intention-to-treat*</u> | 0.02 | -0.14, 0.18 | 0.791 |
| <u>*As-treated*</u> | 0.02 | -0.16, 0.20 | 0.800 |

Abbreviations: ECog-II, revised Everyday Cognition scale.
Notes: Models adjusted for age, sex, education and recreational physical activity. p values represent statistical significance of the group x timepoint interaction term.

## Discussion

The present study examined a 3-month, self-administered, web-based cognitive training programme in middle-aged adults with MetS who exhibited mild cognitive deficits. Participants (mean age 57.8 years, range 40–72) were instructed to complete 24 sessions, two per week, each lasting 45 minutes, at home. While participants were able to access the programme without difficulty, adherence was modest. Only half of the participants completed at least 8 sessions of the cognitive training programme. These observations indicate that home-based cognitive training is feasible in this population, although additional strategies to enhance engagement are needed.

Overall, this low intensity cognitive training was not associated with improvements in the intervention group compared with the control group on any neuropsychological test or in everyday functioning at 3-month or at 12-month follow-up. While small within-group enhancements were observed in both groups on some memory tests, semantic verbal fluency and reaction times of the computerized neuropsychological tests, between-group differences were generally not statistically significant with wide confidence intervals.

Effect estimates occasionally favoured the intervention group (i.e, semantic verbal fluency, Complex Figure-immediate and delay recall, HVLT-R - learning), but none of them reached statistical significance. Fewer effect estimates favoured the control group (i.e., MoCA, Complex Figure-copy), but these associations were inconsistent. As-treated analyses, including only adherent participants, revealed larger effect sizes, particularly in memory tasks, that yet remained statistically non-significant. Performance on computerized neuropsychological tests changed little in both groups using both statistical approaches.

The findings of the present study align with prior investigations using the BrainHQ in individuals with MCI and a mean age above 70 years, in which training-related effects appeared to be limited. ^55–58^. Although statistically significant training effects were not consistently observed, most studies reported effect sizes that favoured the intervention group. The modest magnitude of findings was frequently attributed to limited statistical power, given the relatively small sample sizes (ranging from 17 to 113 participants).

In contrast, other studies have found significant cognitive benefits following computerized cognitive training interventions in individuals with MCI ^59–64^. In these studies, intervention groups presented with cognitive improvements, especially in the trained domains, when compared with active or passive control groups. Although these studies involved participants with comparable demographic profiles and training programme characteristics, the interventions were more frequently supervised by a certified trainer and less often home-based. It is noteworthy that upon meta-analytic integration of these and other similar studies, statistically significant, but small to moderate, gains were noted for cognitive function in older adults with MCI ^30–32,65–67^.

The lack of significant improvement observed in the present study may additionally indicate that the cognitive deficits exhibited by participants with MetS were too subtle to be effectively addressed by the specific training protocol employed. Even though previous studies support cognitive benefits in healthy adults ^24–26,68^, individuals with more pronounced cognitive deficits appear to derive greater benefit from cognitive training programmes ^69–71^.

It is also possible that cognitive training is more effective when combined with other interventions, such as physical exercise alone ^72–75^ or in combination with nutritional intervention ^76–78^. Alongside this, although home-based cognitive training can yield benefits, prior meta-analyses have reported that supervised, group-based cognitive training programmes ^26,67,79^ were apt to display an improvement in cognitive function.

Finally, participant adherence may play a critical role in determining the efficacy of cognitive training interventions. A recent meta-analysis reported that intervention completion rates exceeding 60% are required for older adults with normal cognition to achieve significant cognitive training gains ^80^. In the present study, adherence reached 50%, which may partly explain the absence of significant training effects. Given that the LLM Care protocol requires a minimum of 24 sessions, participants’ completion of only about 15 of the intended sessions suggests limited adherence, and consequently, the results should be interpreted with caution. Consistent with this interpretation, greater cumulative training exposure was generally associated with higher cognitive performance in our study, and higher training exposure was more frequently observed among older participants.

To the best of our knowledge, this is the first study to evaluate the feasibility and efficacy of a short-term, self-administered, computerized cognitive training programme targeting mild cognitive deficits in individuals with MetS, a population group at high risk for developing cognitive dysfunction. Another important feature of this study is the use of a sample drawn from a population-based cohort, selected based on standardized exposure measurements, and monitored over a sufficiently long follow-up period of 1 year. Moreover, cognitive function was assessed using alternate versions of widely administered neuropsychological tests to control for practice effects, and encompassing a broad range of cognitive domains, rather than focusing solely on those targeted by the cognitive training programme. Lastly, cognitive training was delivered through a broadly adopted web-based platform, designed to be user-friendly and to incorporate adaptive difficulty levels, without positing strict timetabling constraints.

However, important limitations need to be acknowledged. First, the small sample size might have limited the ability of the cognitive programme to detect statistically significant changes on neuropsychological tests. Furthermore, although most participants were recruited from the comprehensive EHS database (n = 24), a smaller group was enrolled based on self-referral (n = 12). Second, half of the participants were non-compliant with the training protocol, which probably influenced the outcome of the study. One conceivable explanation is that younger participants with subtle cognitive deficits have been less motivated to persist with the cognitive training programme due to a lower perceived benefit.

## Conclusions

Despite these limitations, the current study contributes to the growing body of literature regarding the impact of cognitive training programmes on individuals’ cognitive function. Short-term, self-administered programmes, such as the one examined, may represent a cost-efficient approach for supporting cognitive health, although their benefits appear limited under conditions of low adherence. Clinical trials with larger sample sizes, extended intervention durations and longer follow-up periods are warranted. Future studies should also incorporate more rigorous adherence monitoring, potentially through clinician-delivered or supervised interventions to enhance acceptability, engagement and ensure protocol fidelity.

## Data availability

The datasets used and analyzed during the current study are available from the corresponding author on reasonable request.

## Supporting information

Summplementary Table

## Authors’ contributions

MK, EA and KKT Research conception and design; MK Data collection and drafting of manuscript; MK and GM Data analysis; AK, YT, AV, VZ, ER, TH and EN Review & editing. All authors revised and approved the final manuscript.

## Ethics declarations

The study was approved by the Research Ethics Committee of the University of Ioannina (Approval number 27819/25-05-2022), and was conducted in accordance with the Declaration of Helsinki. All participants provided written informed consent prior to participation in the study.

## References

1 González-Castañeda, H. et al. Neuropsychology of metabolic syndrome: A systematic review and meta-analysis. Cogent Psychology 8, 1913878 (2021).

2 Siervo, M., Harrison, S. L., Jagger, C., Robinson, L. & Stephan, B. C. Metabolic syndrome and longitudinal changes in cognitive function: a systematic review and meta-analysis. J Alzheimers Dis 41, 151–161, doi:10.3233/JAD-132279 (2014).

3 Kazlauskaite, R. et al. Is Midlife Metabolic Syndrome Associated With Cognitive Function Change? The Study of Women’s Health Across the Nation. The Journal of clinical endocrinology and metabolism 105, e1093–1105, doi:10.1210/clinem/dgaa067 (2020).

4 Reijmer, Y. D. et al. The metabolic syndrome, atherosclerosis and cognitive functioning in a non-demented population: the Hoorn Study. Atherosclerosis 219, 839–845, doi:10.1016/j.atherosclerosis.2011.08.032 (2011).

5 Wooten, T. et al. Metabolic risk in older adults is associated with impaired sustained attention. Neuropsychology 33, 947–955, doi:10.1037/neu0000554 (2019).

6 Falkowski, J., Atchison, T., Debutte-Smith, M., Weiner, M. F. & O’Bryant, S. Executive functioning and the metabolic syndrome: a project FRONTIER study. Archives of clinical neuropsychology : the official journal of the National Academy of Neuropsychologists 29, 47–53, doi:10.1093/arclin/act078 (2014).

7 Rouch, I. et al. Metabolic syndrome is associated with poor memory and executive performance in elderly community residents: the PROOF study. The American journal of geriatric psychiatry : official journal of the American Association for Geriatric Psychiatry 22, 1096–1104, doi:10.1016/j.jagp.2014.01.005 (2014).

8 Raffaitin, C. et al. Metabolic syndrome and cognitive decline in French elders: the Three-City Study. Neurology 76, 518–525, doi:10.1212/WNL.0b013e31820b7656 (2011).

9 Foret, J. T. et al. Metabolic Syndrome and Cognitive Function in Midlife. Archives of clinical neuropsychology : the official journal of the National Academy of Neuropsychologists 36, 897–907, doi:10.1093/arclin/acaa112 (2021).

10 Hassenstab, J. J., Sweat, V., Bruehl, H. & Convit, A. Metabolic syndrome is associated with learning and recall impairment in middle age. Dementia and geriatric cognitive disorders 29, 356–362, doi:10.1159/000296071 (2010).

11 González, H. M. et al. Metabolic Syndrome and Neurocognition Among Diverse Middle-Aged and Older Hispanics/Latinos: HCHS/SOL Results. Diabetes care 41, 1501–1509, doi:10.2337/dc17-1896 (2018).

12 Luo, L., Yang, M., Hao, Q., Yue, J. & Dong, B. Cross-sectional study examining the association between metabolic syndrome and cognitive function among the oldest old. Journal of the American Medical Directors Association 14, 105–108, doi:10.1016/j.jamda.2012.10.001 (2013).

13 Liu, C. L. et al. Late-life metabolic syndrome prevents cognitive decline among older men aged 75 years and over: one-year prospective cohort study. The journal of nutrition, health & aging 17, 523–526, doi:10.1007/s12603-013-0010-2 (2013).

14 Jekel, K. et al. Mild cognitive impairment and deficits in instrumental activities of daily living: a systematic review. Alzheimers Res Ther 7, 17, doi:10.1186/s13195-015-0099-0 (2015).

15 Lindbergh, C. A., Dishman, R. K. & Miller, L. S. Functional Disability in Mild Cognitive Impairment: A Systematic Review and Meta-Analysis. Neuropsychol Rev 26, 129–159, doi:10.1007/s11065-016-9321-5 (2016).

16 Reppermund, S. et al. The relationship of neuropsychological function to instrumental activities of daily living in mild cognitive impairment. Int J Geriatr Psychiatry 26, 843–852, doi:10.1002/gps.2612 (2011).

17 Roberts, R. O. et al. Higher risk of progression to dementia in mild cognitive impairment cases who revert to normal. Neurology 82, 317–325, doi:10.1212/WNL.0000000000000055 (2014).

18 Chen, Y. et al. Progression from normal cognition to mild cognitive impairment in a diverse clinic-based and community-based elderly cohort. Alzheimers Dement 13, 399–405, doi:10.1016/j.jalz.2016.07.151 (2017).

19 Blacker, D. et al. Neuropsychological measures in normal individuals that predict subsequent cognitive decline. Arch Neurol 64, 862–871, doi:10.1001/archneur.64.6.862 (2007).

20 Knopman, D. S. et al. Midlife vascular risk factors and midlife cognitive status in relation to prevalence of mild cognitive impairment and dementia in later life: The Atherosclerosis Risk in Communities Study. Alzheimers Dement 14, 1406–1415, doi:10.1016/j.jalz.2018.03.011 (2018).

21 Ng, T. P. et al. Metabolic Syndrome and the Risk of Mild Cognitive Impairment and Progression to Dementia: Follow-up of the Singapore Longitudinal Ageing Study Cohort. JAMA Neurol 73, 456–463, doi:10.1001/jamaneurol.2015.4899 (2016).

22 Qiu, S. D. et al. Associations of metabolic syndrome with risks of dementia and cognitive impairment: A systematic review and meta-analysis. J Alzheimers Dis 105, 15–27, doi:10.1177/13872877251326553 (2025).

23 Reddy, K. J. in Innovations in Neurocognitive Rehabilitation: Harnessing Technology for Effective Therapy 171–209 (Springer, 2025).

24 Chiu, H. L. et al. The effect of cognitive-based training for the healthy older people: A meta-analysis of randomized controlled trials. PLoS One 12, e0176742, doi:10.1371/journal.pone.0176742 (2017).

25 Butler, M. et al. Does Cognitive Training Prevent Cognitive Decline?: A Systematic Review. Ann Intern Med 168, 63–68, doi:10.7326/M17-1531 (2018).

26 Kelly, M. E. et al. The impact of cognitive training and mental stimulation on cognitive and everyday functioning of healthy older adults: a systematic review and meta-analysis. Ageing Res Rev 15, 28–43, doi:10.1016/j.arr.2014.02.004 (2014).

27 Zoupa, E. et al. Cognitive Rehabilitation in Schizophrenia-Associated Cognitive Impairment: A Review. Neurol Int 15, 12–23, doi:10.3390/neurolint15010002 (2022).

28 Woolf, C. et al. A Systematic Review and Meta-Analysis of Cognitive Training in Adults with Major Depressive Disorder. Neuropsychol Rev 32, 419–437, doi:10.1007/s11065-021-09487-3 (2022).

29 Cella, M. et al. Cognitive remediation for inpatients with psychosis: a systematic review and meta-analysis. Psychol Med 50, 1062–1076, doi:10.1017/S0033291720000872 (2020).

30 Zhang, H. et al. Effect of computerised cognitive training on cognitive outcomes in mild cognitive impairment: a systematic review and meta-analysis. BMJ Open 9, e027062, doi:10.1136/bmjopen-2018-027062 (2019).

31 Hill, N. T. et al. Computerized Cognitive Training in Older Adults With Mild Cognitive Impairment or Dementia: A Systematic Review and Meta-Analysis. Am J Psychiatry 174, 329–340, doi:10.1176/appi.ajp.2016.16030360 (2017).

32 Sherman, D. S., Mauser, J., Nuno, M. & Sherzai, D. The Efficacy of Cognitive Intervention in Mild Cognitive Impairment (MCI): a Meta-Analysis of Outcomes on Neuropsychological Measures. Neuropsychol Rev 27, 440–484, doi:10.1007/s11065-017-9363-3 (2017).

33 Bahar-Fuchs, A., Martyr, A., Goh, A. M., Sabates, J. & Clare, L. Cognitive training for people with mild to moderate dementia. Cochrane Database Syst Rev 3, CD013069, doi:10.1002/14651858.CD013069.pub2 (2019).

34 Lampit, A. et al. Computerized cognitive training in multiple sclerosis: a systematic review and meta-analysis. Neurorehabilitation and Neural Repair 33, 695–706 (2019).

35 Fernandes, H. A., Richard, N. M. & Edelstein, K. Cognitive rehabilitation for cancer-related cognitive dysfunction: a systematic review. Support Care Cancer 27, 3253–3279, doi:10.1007/s00520-019-04866-2 (2019).

36 Rogers, J. M., Foord, R., Stolwyk, R. J., Wong, D. & Wilson, P. H. General and Domain-Specific Effectiveness of Cognitive Remediation after Stroke: Systematic Literature Review and Meta-Analysis. Neuropsychol Rev 28, 285–309, doi:10.1007/s11065-018-9378-4 (2018).

37 Noubiap, J. J. et al. Geographic distribution of metabolic syndrome and its components in the general adult population: A meta-analysis of global data from 28 million individuals. Diabetes Res Clin Pract 188, 109924, doi:10.1016/j.diabres.2022.109924 (2022).

38 Kanellopoulou, A. et al. Awareness, knowledge and trust in the Greek authorities towards COVID-19 pandemic: results from the Epirus Health Study cohort. BMC Public Health 21, 1125, doi:10.1186/s12889-021-11193-x (2021).

39 Koutsonida, M. et al. Metabolic syndrome and cognitive deficits in the Greek cohort of Epirus Health Study. Neurol Sci 44, 3523–3533, doi:10.1007/s10072-023-06835-4 (2023).

40 Vlachou, C. & Kosmidis, Μ. The Greek Trail Making Test: Preliminary normative data for clinical and research use. Psychology: the Journal of the Hellenic Psychological Society 9, 336–352, doi:10.12681/psy_hps.24068 (2020).

41 Kosmidis, M. H., Vlahou, C. H., Panagiotaki, P. & Kiosseoglou, G. The verbal fluency task in the Greek population: normative data, and clustering and switching strategies. Journal of the International Neuropsychological Society : JINS 10, 164–172, doi:10.1017/s1355617704102014 (2004).

42 Kokkinis, N., Kosmidis, M. H., Kiosseoglou, G. & Aretouli, E. Neuropsychological Assessment Battery (SyNePsy): Normative data and investigation of diagnostic validity in mixed neurological population, Aristotle University of Thessaloniki, (2019).

43 Whitehead, A. L., Julious, S. A., Cooper, C. L. & Campbell, M. J. Estimating the sample size for a pilot randomised trial to minimise the overall trial sample size for the external pilot and main trial for a continuous outcome variable. Stat Methods Med Res 25, 1057–1073, doi:10.1177/0962280215588241 (2016).

44 Alberti, K. G., Zimmet, P. & Shaw, J. The metabolic syndrome--a new worldwide definition. Lancet (London, England) 366, 1059–1062, doi:10.1016/s0140-6736(05)67402-8 (2005).

45 Grundy, S. M. et al. Diagnosis and management of the metabolic syndrome: an American Heart Association/National Heart, Lung, and Blood Institute Scientific Statement. Circulation 112, 2735–2752, doi:10.1161/circulationaha.105.169404 (2005).

46 Petersen, R. C. Mild cognitive impairment as a diagnostic entity. J Intern Med 256, 183–194, doi:10.1111/j.1365-2796.2004.01388.x (2004).

47 in Risk Reduction of Cognitive Decline and Dementia: WHO Guidelines WHO Guidelines Approved by the Guidelines Review Committee (2019).

48 Bamidis, P., et al. (2012).

49 Nasreddine, Z. S. et al. The Montreal Cognitive Assessment, MoCA: a brief screening tool for mild cognitive impairment. J Am Geriatr Soc 53, 695–699, doi:10.1111/j.1532-5415.2005.53221.x (2005).

50 Benedict, R. H., Schretlen, D., Groninger, L. & Brandt, J. Hopkins Verbal Learning Test–Revised: Normative data and analysis of inter-form and test-retest reliability. The Clinical Neuropsychologist 12, 43–55 (1998).

51 Taylor, L. B. Localisation of cerebral lesions by psychological testing. Clin Neurosurg 16, 269–287, doi:10.1093/neurosurgery/16.cn_suppl_1.269 (1969).

52 Loring, D. & Meador, K. The Medical College of Georgia (MCG) complex figures: four forms for follow-up. Rey-Osterrieth handbook. Psychological Assessment Resources (2003).

53 Kramer, J. H. et al. NIH EXAMINER: conceptualization and development of an executive function battery. Journal of the International Neuropsychological Society : JINS 20, 11–19, doi:10.1017/S1355617713001094 (2014).

54 Farias, S. T. et al. The Measurement of Everyday Cognition (ECog): Revisions and Updates. Alzheimer Dis Assoc Disord 35, 258–264, doi:10.1097/WAD.0000000000000450 (2021).

55 Duff, K. et al. Computerized Cognitive Training in Amnestic Mild Cognitive Impairment: A Randomized Clinical Trial. J Geriatr Psychiatry Neurol 35, 400–409, doi:10.1177/08919887211006472 (2022).

56 Styliadis, C., Kartsidis, P., Paraskevopoulos, E., Ioannides, A. A. & Bamidis, P. D. Neuroplastic effects of combined computerized physical and cognitive training in elderly individuals at risk for dementia: an eLORETA controlled study on resting states. Neural Plast 2015, 172192, doi:10.1155/2015/172192 (2015).

57 Knoefel, F. et al. Implementation of a Brain Training Pilot Study For People With Mild Cognitive Impairment. Can Geriatr J 21, 264–268, doi:10.5770/cgj.21.304 (2018).

58 Barnes, D. E. et al. Computer-based cognitive training for mild cognitive impairment: results from a pilot randomized, controlled trial. Alzheimer Dis Assoc Disord 23, 205–210, doi:10.1097/WAD.0b013e31819c6137 (2009).

59 Herrera, C., Chambon, C., Michel, B. F., Paban, V. & Alescio-Lautier, B. Positive effects of computer-based cognitive training in adults with mild cognitive impairment. Neuropsychologia 50, 1871–1881, doi:10.1016/j.neuropsychologia.2012.04.012 (2012).

60 Lin, F. et al. Cognitive and Neural Effects of Vision-Based Speed-of-Processing Training in Older Adults with Amnestic Mild Cognitive Impairment: A Pilot Study. J Am Geriatr Soc 64, 1293–1298, doi:10.1111/jgs.14132 (2016).

61 Wu, J. et al. Computerized Cognitive Training Enhances Episodic Memory by Down-Modulating Posterior Cingulate-Precuneus Connectivity in Older Persons With Mild Cognitive Impairment: A Randomized Controlled Trial. The American journal of geriatric psychiatry : official journal of the American Association for Geriatric Psychiatry 31, 820–832, doi:10.1016/j.jagp.2023.04.008 (2023).

62 Valdes, E. G., O’Connor, M. L. & Edwards, J. D. The effects of cognitive speed of processing training among older adults with psychometrically- defined mild cognitive impairment. Curr Alzheimer Res 9, 999–1009, doi:10.2174/156720512803568984 (2012).

63 Nousia, A. et al. Beneficial effect of computer-based multidomain cognitive training in patients with mild cognitive impairment. Appl Neuropsychol Adult 28, 717–726, doi:10.1080/23279095.2019.1692842 (2021).

64 Damirchi, A., Hosseini, F. & Babaei, P. Mental Training Enhances Cognitive Function and BDNF More Than Either Physical or Combined Training in Elderly Women With MCI: A Small-Scale Study. Am J Alzheimers Dis Other Demen 33, 20–29, doi:10.1177/1533317517727068 (2018).

65 Li, R., Geng, J., Yang, R., Ge, Y. & Hesketh, T. Effectiveness of Computerized Cognitive Training in Delaying Cognitive Function Decline in People With Mild Cognitive Impairment: Systematic Review and Meta-analysis. J Med Internet Res 24, e38624, doi:10.2196/38624 (2022).

66 Xu, Z. et al. Comparative Effectiveness of Interventions for Global Cognition in Patients With Mild Cognitive Impairment: A Systematic Review and Network Meta-Analysis of Randomized Controlled Trials. Front Aging Neurosci 13, 653340, doi:10.3389/fnagi.2021.653340 (2021).

67 Chan, A. T. C., Ip, R. T. F., Tran, J. Y. S., Chan, J. Y. C. & Tsoi, K. K. F. Computerized cognitive training for memory functions in mild cognitive impairment or dementia: a systematic review and meta-analysis. NPJ Digit Med 7, 1, doi:10.1038/s41746-023-00987-5 (2024).

68 Bonnechere, B., Langley, C. & Sahakian, B. J. The use of commercial computerised cognitive games in older adults: a meta-analysis. Sci Rep 10, 15276, doi:10.1038/s41598-020-72281-3 (2020).

69 Traut, H. J., Guild, R. M. & Munakata, Y. Why Does Cognitive Training Yield Inconsistent Benefits? A Meta-Analysis of Individual Differences in Baseline Cognitive Abilities and Training Outcomes. Front Psychol 12, 662139, doi:10.3389/fpsyg.2021.662139 (2021).

70 Roheger, M., Kalbe, E., Corbett, A., Brooker, H. & Ballard, C. Lower cognitive baseline scores predict cognitive training success after 6 months in healthy older adults: Results of an online RCT. Int J Geriatr Psychiatry 35, 1000–1008, doi:10.1002/gps.5322 (2020).

71 Maraver, M. J., Gomez-Ariza, C. J., Borella, E. & Bajo, M. T. Baseline capacities and motivation in executive control training of healthy older adults. Aging Ment Health 26, 595–603, doi:10.1080/13607863.2020.1858755 (2022).

72 Montero-Odasso, M. et al. Effects of Exercise Alone or Combined With Cognitive Training and Vitamin D Supplementation to Improve Cognition in Adults With Mild Cognitive Impairment: A Randomized Clinical Trial. JAMA Netw Open 6, e2324465, doi:10.1001/jamanetworkopen.2023.24465 (2023).

73 Donnezan, L. C., Perrot, A., Belleville, S., Bloch, F. & Kemoun, G. Effects of simultaneous aerobic and cognitive training on executive functions, cardiovascular fitness and functional abilities in older adults with mild cognitive impairment. Mental Health and Physical Activity 15, 78–87 (2018).

74 Castellote-Caballero, Y., Carcelen Fraile, M. D. C., Aibar-Almazan, A., Afanador-Restrepo, D. F. & Gonzalez-Martin, A. M. Effect of combined physical-cognitive training on the functional and cognitive capacity of older people with mild cognitive impairment: a randomized controlled trial. BMC Med 22, 281, doi:10.1186/s12916-024-03469-x (2024).

75 Bamidis, P. D. et al. Gains in cognition through combined cognitive and physical training: the role of training dosage and severity of neurocognitive disorder. Front Aging Neurosci 7, 152, doi:10.3389/fnagi.2015.00152 (2015).

76 Wang, P. et al. Effects of a Multicomponent Intervention With Cognitive Training and Lifestyle Guidance for Older Adults at Risk of Dementia: A Randomized Controlled Trial. J Clin Psychiatry 85, doi:10.4088/JCP.23m15112 (2024).

77 Ngandu, T. et al. A 2 year multidomain intervention of diet, exercise, cognitive training, and vascular risk monitoring versus control to prevent cognitive decline in at-risk elderly people (FINGER): a randomised controlled trial. Lancet (London, England) 385, 2255–2263, doi:10.1016/S0140-6736(15)60461-5 (2015).

78 Chatterjee, P. et al. Effect of Multimodal Intervention (computer based cognitive training, diet and exercise) in comparison to health awareness among older adults with Subjective Cognitive Impairment (MISCI-Trial)-A Pilot Randomized Control Trial. PLoS One 17, e0276986, doi:10.1371/journal.pone.0276986 (2022).

79 Lampit, A., Hallock, H. & Valenzuela, M. Computerized cognitive training in cognitively healthy older adults: a systematic review and meta-analysis of effect modifiers. PLoS Med 11, e1001756, doi:10.1371/journal.pmed.1001756 (2014).

80 Li, Z., He, H., Chen, Y. & Guan, Q. Effects of engagement, persistence and adherence on cognitive training outcomes in older adults with and without cognitive impairment: a systematic review and meta-analysis of randomised controlled trials. Age Ageing 53, doi:10.1093/ageing/afad247 (2024).

