## Supplementary material for "Self-administered computerized cognitive training for cognitive deficits in individuals with metabolic syndrome: a randomized controlled trial": Summplementary Table

**Supplementary Table S1.** Detailed description of BrainHQ tasks incorporated into the cognitive training programme.

| Tasks | Description | Cognitive domain |
|---|---|---|
| *People skills* | | |
| **Face Facts** | A series of profile pictures are shown, each accompanied by facts about the individuals' lives. Then, one person's image is shown again, and the task is to select the correct fact about them from a list of options.<br>The task adapts by increasing the number of people, making facts more similar and changing genders. | Memory |
| **In the Know** | Three characters engage in a conversation, making statements about people they know. Following the conversation, a questionnaire is presented, and participants must select the correct answers.<br>The task adapts by increasing the speed at which the characters speak and making sentences more complex. | Memory |
| *Navigation* | | |
| **Mental Map** | A grid appears with several objects placed at different positions. The images then disappear, and the grid may be rotated, flipped, or moved. The task is to drag the missing objects back to their correct positions on the grid, taking into account the new orientation.<br>The task adapts by increasing the complexity of grid movements and presenting more objects within the grid. | Spatial memory |
| **True North** | Instructions are given to take a specific train in a particular direction to reach a new station. The task is to identify the correct train based on a compass display. At each subsequent station, the orientation may change, requiring to reorient and remember the next set of instructions.<br>The task adapts by increasing the complexity of both the instructions and the orientation information. | Working memory |
| **Optic Flow** | A target shape is displayed on a sign along a driving route. The task is to identify matching shapes on approaching vehicles and roadside objects.<br>The task adapts by making the shapes more similar, changing the driving conditions (ex. raining), and increasing the complexity of the background. | Attention |
| *Intelligence* | | |
| **Auditory Ace** | Information about playing cards is presented auditory, one card at a time. The task is to decide whether the current card matches the one presented a specific number of steps earlier in the sequence.<br>The task adapts by increasing the number of items to remember in the sequence and presenting the information at a faster pace. | Working memory |
| **Card Shark** | Playing cards are presented one by one and added to a sequence, but each card is turned face down after being | Working memory |

| | | |
|---|---|---|
| | shown. The task is to decide whether the current card matches the one shown a specific number of steps earlier in the sequence.<br>The task adapts by increasing the number of items to remember and presenting the information at a faster pace. | |
| ***Attention*** | | |
| **Divided Attention** | The task is to quickly determine whether two side-by-side shapes match a predefined criterion (ex. same color, shape, filling) within a limited time.<br>The task adapts by reducing the available time and introducing multiple criteria. | Attention |
| **Mixed Signals** | A criterion is displayed at the top of the screen — for example, matching a spoken number to the number of items shown. If the spoken word is matched with the image according to the given criterion, a response is required. If there is no match, no action should be taken.<br>The task adapts as the target becomes progressively more difficult to discern. | Attention |
| ***Brain speed*** | | |
| **Sound Sweeps** | Sounds that either rise or fall in pitch are presented, and the task is to determine the direction—up or down.<br>The task adapts by presenting the information at a faster pace. | Processing speed |

**Supplementary Table S2.** Baseline socio-demographic characteristics of enrolled and non-enrolled study participants.

| Characteristics | Enrolled (n=36) | Not enrolled (n=50) | p value |
|---|---|---|---|
| **Age**, years | 57.13 ± 8.29 | 56.37 ± 6.91 | 0.643 [a] |
| **Sex** | | | 0.967 [b] |
| Female | 20 (55.56%) | 28 (56.00%) | |
| Men | 16 (44.44%) | 22 (44.00%) | |
| **Education** | | | 0.765 [b] |
| Primary and secondary school* | 6 (16.67%) | 7 (14.00%) | |
| High school** | 7 (19.44%) | 13 (26.00%) | |
| Higher education*** | 23 (63.89%) | 30 (60.00%) | |
| **MetS components** | | | |
| Obesity | 30 (83.33%) | 33 (66.00%) | 0.096 [b] |
| Dyslipidemia | 30 (83.33%) | 33 (66.00%) | 0.073 [b] |
| Hypertension | 30 (83.33%) | 27 (56.25%) | 0.009 [b] |
| Hyperglycemia | 13 (36.11%) | 8 (16.33%) | 0.037 [b] |
| **Physical activity**, MET-hours/week | 7.82 ± 12.01 | 12.03 ± 16.57 | 0.198 [a] |
| **Smoking** | | | 0.327 [b] |
| Never | 21 (58.33%) | 21 (42.00%) | |
| Former | 4 (11.11%) | 4 (16.00%) | |
| Current | 11 (30.56%) | 21 (42.00%) | |
| **Alcohol consumption** | | | 0.959 [b] |
| Never | 19 (52.78%) | 24 (48.00%) | |
| Monthly | 4 (11.11%) | 5 (10.00%) | |
| Weekly | 10 (27.78%) | 16 (32.00%) | |
| Almost everyday | 3 (8.33%) | 5 (10.00%) | |
| **Income** | | | 0.389 [b] |
| Low [1] | 5 (13.89%) | 7 (15.91%) | |
| Average [2] | 5 (13.89%) | 10 (22.73%) | |
| High [3] | 9 (25.00%) | 5 (11.36%) | |
| Very high [4] | 17 (47.22%) | 22 (50.00%) | |

Abbreviations: MetS, Metabolic Syndrome; MET, metabolic equivalents of energy expenditure.

Notes: *Elementary school or junior high school, up to 9 years of education. **High school, up to 12 years of education. ***University degree/MSc/PhD/Postdoc, more than 13 years of education.

[1] up to 900 euros, [2] 901-1400 euros, [3] 1401-2000 euros, [4] >2000 euros.

[a] Comparisons using t-test. [b] Comparisons using $x^2$ test. Mean ± standard deviation and frequency (percentage) are presented for continuous and categorical variables, respectively.

**Supplementary Table S3.** Baseline socio-demographic characteristics and cognitive scores of lost study participants.

| Characteristics | Completed (n=32) | Lost (n=4) | p value |
|---|---|---|---|
| **Age**, years | 57.82 ± 8.22 | 51.62 ± 7.67 | 0.161 [a] |
| **Sex** | | | 1.000 [b] |
| Female | 18 (56.25%) | 2 (50.00%) | |
| Men | 14 (43.75%) | 2 (50.00%) | |
| **Education** | | | 0.429 [b] |
| Primary and secondary school* | 6 (18.75%) | 0 (0.00%) | |
| High school** | 7 (21.88%) | 0 (0.00%) | |
| Higher education*** | 19 (59.38%) | 4 (100.00%) | |
| **MetS components** | | | |
| Obesity | 27 (84.38%) | 3 (75.00%) | 1.000 [b] |
| Dyslipidemia | 28 (87.50%) | 2 (50.00%) | 0.121 [b] |
| Hypertension | 26 (81.25%) | 4 (100.00%) | 1.000 [b] |
| Hyperglycemia | 11 (31.58%) | 2 (50.00%) | 0.609 [b] |
| **Physical activity**, MET-hours/week | 7.77 ± 12.12 | 8.25 ± 12.82 | 0.941 [a] |
| **Smoking** | | | 0.207 [b] |
| Never | 20 (62.50%) | 1 (25.00%) | |
| Former | 3 (9.38%) | 1 (25.00%) | |
| Current | 9 (28.13%) | 2 (50.00%) | |
| **Alcohol consumption** | | | 0.219 [b] |
| Never | 18 (56.25%) | 1 (25.00%) | |
| Monthly | 3 (9.38%) | 1 (25.00%) | |
| Weekly | 9 (28.13%) | 1 (25.00%) | |

| Neuropsychological test | Completed (n=32) | Lost (n=4) | p value |
|---|---|---|---|
| *Paper-and-pencil* | | | |
| **MoCA** | 24.75 ± 3.17 | 27.00 ± 2.58 | 0.184 [a] |
| **Taylor complex figure** | | | |
| Copy | 34.78 ± 1.70 | 34.00 ± 2.16 | 0.404 [a] |
| Immediate recall | 25.30 ± 6.38 | 23.38 ± 3.50 | 0.562 [a] |
| Delayed recall | 23.70 ± 7.73 | 22.63 ± 6.34 | 0.791 [a] |
| **Hopkins Verbal Learning Test** | | | |
| Learning | 22.69 ± 4.40 | 23.50 ± 3.11 | 0.724 [a] |
| Recall | 8.22 ± 1.95 | 8.50 ± 2.08 | 0.788 [a] |
| Recognition | 21.53 ± 1.34 | 22.75 ± 1.26 | 0.095 [a] |
| **Verbal fluency** | | | |
| Phonemic | 11.25 ± 3.58 | 11.25 ± 3.59 | 1.000 [a] |
| Semantic | 15.75 ± 3.18 | 12.75 ± 5.32 | 0.108 [a] |
| **Digit span** | | | |
| Forward | 6.66 ± 2.12 | 7.75 ± 0.96 | 0.320 [a] |
| Backward | 5.38 ± 2.21 | 6.75 ± 1.26 | 0.235 [a] |
| *Computerized* | | | |
| **Continuous Performance Test** | | | |
| Total correct | 97.75 ± 2.06 | 98.50 ± 1.00 | 0.483 [a] |
| Mean time (ms) | 0.52 ± 0.07 | 0.50 ± 0.05 | 0.524 [a] |
| **Flanker task** | | | |
| Total correct | 47.56 ± 0.72 | 48.00 ± 0.00 | 0.236 [a] |

| | | | | | | | |
|---|---|---|---|---|---|---|---|
| Almost everyday | 2 (6.25%) | 1 (25.00%) | | Mean time (ms) | 1.00 ± 0.36 | 0.79 ± 0.22 | 0.273 [a] |
| **Income** | | | 1.000 [b] | **Set-Shifting task** | | | |
| Low [1] | 5 (15.63%) | 0 (0.00%) | | Total correct | 101.03 ± 3.00 | 102.75 ± 1.89 | 0.275 [a] |
| Average [2] | 5 (15.63%) | 0 (0.00%) | | Mean time (ms) | 1.00 ± 0.32 | 0.82 ± 0.24 | 0.274 [a] |
| High [3] | 8 (25.00%) | 1 (25.00%) | | **N-Back task** | 26.44 ± 2.34 | 25.25 ± 4.27 | 0.390 [a] |
| Very high [4] | 14 (43.75%) | 3 (75.00%) | | *Functional abilities* | | | |
| | | | | **ECog-II** | 1.19 ± 0.18 | 1.26 ± 0.80 | 0.460 [a] |

Abbreviations: MetS, Metabolic Syndrome; MET, metabolic equivalents of energy expenditure; MoCA, Montreal Cognitive Assessment; ms, milliseconds; ECog-II, revised Everyday Cognition scale.

Notes: *Elementary school or junior high school, up to 9 years of education. **High school, up to 12 years of education. ***University degree/MSc/PhD/Postdoc, more than 13 years of education.

[1] up to 900 euros, [2] 901-1400 euros, [3] 1401-2000 euros, [4] >2000 euros.

[a] Comparisons using t-test. [b] Comparisons using Fisher's exact test. Mean ± standard deviation and frequency (percentage) are presented for continuous and categorical variables, respectively.

**Supplementary Table S4.** Baseline socio-demographic characteristics and cognitive scores of study participants according to as-treated principle.

| Characteristics | Group | | p value |
|---|---|---|---|
| | Cognitive training (n=9) | Control (n=24) | |
| **Age**, years | 58.09 ± 5.63 | 57.73 ± 8.94 | 0.910 [a] |
| **Sex** | | | 0.698 [b] |
| Female | 6 (66.67%) | 13 (54.17%) | |
| Men | 3 (33.33%) | 11 (45.83%) | |
| **Education** | | | 0.763 [b] |
| Primary and secondary school* | 1 (11.11%) | 5 (20.83%) | |
| High school** | 2 (22.22%) | 5 (20.83%) | |
| Higher education*** | 6 (66.67%) | 14 (58.33%) | |
| **MetS components** | | | |
| Obesity | 9 (100.00%) | 19 (79.17%) | 0.290 [b] |
| Dyslipidemia | 9 (100.00%) | 20 (83.33%) | 0.555 [b] |
| Hypertension | 8 (88.89%) | 19 (79.17%) | 1.000 [b] |
| Hyperglycemia | 3 (33.33%) | 8 (33.33%) | 1.000 [b] |
| **Physical activity**, MET-hours/week | 14.39 ± 21.11 | 6.08 ± 6.24 | 0.086 [a] |
| **Smoking** | | | 0.855 [b] |
| Never | 6 (66.67%) | 14 (58.33%) | |
| Former | 1 (11.11%) | 2 (8.33%) | |
| Current | 2 (22.22%) | 8 (33.33%) | |
| **Alcohol consumption** | | | 0.928 [b] |
| Never | 4 (44.44%) | 14 (58.33%) | |
| Monthly | 1 (11.11%) | 2 (8.33%) | |

| Neuropsychological test | Group | | p value |
|---|---|---|---|
| | Cognitive training (n=9) | Control (n=24) | |
| *Paper-and-pencil* | | | |
| **MoCA** | 25.78 ± 2.39 | 24.33 ± 3.32 | 0.243 [a] |
| **Taylor complex figure** | | | |
| Copy | 35.33 ± 1.00 | 34.54 ± 1.84 | 0.233 [a] |
| Immediate recall | 25.78 ± 6.59 | 24.90± 6.39 | 0.728 [a] |
| Delayed recall | 23.33 ± 9.85 | 23.46 ± 7.09 | 0.968 [a] |
| **Hopkins Verbal Learning Test** | | | |
| Learning | 23.56 ± 3.91 | 22.25 ± 4.53 | 0.452 [a] |
| Recall | 7.56 ± 2.30 | 8.38 ± 1.81 | 0.291 [a] |
| Recognition | 21.56 ± 1.51 | 21.58 ± 1.32 | 0.959 [a] |
| **Verbal fluency** | | | |
| Phonemic | 11.56 ± 4.56 | 11.21 ± 3.19 | 0.806 [a] |
| Semantic | 16.78 ± 3.23 | 15.50 ± 3.16 | 0.312 [a] |
| **Digit span** | | | |
| Forward | 6.89 ± 1.62 | 6.67 ± 2.32 | 0.794 [a] |
| Backward | 5.33 ± 1.23 | 5.46 ± 2.48 | 0.887 [a] |
| *Computerized* | | | |
| **Continuous Performance Test** | | | |
| Total correct | 98.00 ± 2.45 | 97.71 ± 1.92 | 0.721 [a] |
| Mean time (ms) | 0.54 ± 0.07 | 0.51 ± 0.07 | 0.369 [a] |
| **Flanker task** | | | |

| | | | | | | | |
|---|---|---|---|---|---|---|---|
| Weekly | 3 (33.33%) | 6 (25.00%) | | Total correct | 47.89 ± 0.33 | 47.46 ± 0.78 | 0.122 [a] |
| Almost everyday | 1 (11.11%) | 2 (8.33%) | | Mean time (ms) | 1.07 ± 0.29 | 0.97 ± 0.37 | 0.476 [a] |
| **Income** | | | 0. 155 [b] | **Set-Shifting task** | | | |
| Low [1] | 1 (11.11%) | 4 (16.67%) | | Total correct | 102.56 ± 1.59 | 100.58 ± 3.24 | 0.093 [a] |
| Average [2] | 0 (0.00%) | 5 (20.83%) | | Mean time (ms) | 1.05 ± 0.29 | 0.99 ± 0.33 | 0.654 [a] |
| High [3] | 1 (11.11%) | 7 (29.17%) | | **N-Back task** | 25.78 ± 3.15 | 26.38 ± 2.48 | 0.571 [a] |
| Very high [4] | 7 (77.78%) | 8 (33.33%) | | *Functional abilities* | | | |
| | | | | **ECog-II** | 1.20 ± 0.18 | 1.19 ± 0.19 | 0.805 [a] |

Abbreviations: MetS, Metabolic Syndrome; MET, metabolic equivalents of energy expenditure; MoCA, Montreal Cognitive Assessment; ms, milliseconds; ECog-II, revised Everyday Cognition scale.

Notes: *Elementary school or junior high school, up to 9 years of education. **High school, up to 12 years of education. ***University degree/MSc/PhD/Postdoc, more than 13 years of education.

[1] up to 900 euros, [2] 901-1400 euros, [3] 1401-2000 euros, [4] >2000 euros.

[a] Comparisons using t-test. [b] Comparisons using Fisher's exact test. Mean ± standard deviation and frequency (percentage) are presented for continuous and categorical variables, respectively.

**Supplementary Table S5.** Baseline socio-demographic characteristics and cognitive scores of non-adherent study participants.

| Characteristics | Group | | p value |
|---|---|---|---|
| | Adherent (n=9) | Non-adherent (n=7) | |
| **Age**, years | 58.09 ± 5.63 | 54.39 ± 6.26 | 0.234 [a] |
| **Sex** | | | 1.000 [b] |
| Female | 6 (66.67%) | 4 (57.14%) | |
| Men | 3 (33.33%) | 3 (42.86%) | |
| **Education** | | | 1.000 [b] |
| Primary and secondary school* | 1 (11.11%) | 0 (0.00%) | |
| High school** | 2 (22.22%) | 1 (14.29%) | |
| Higher education*** | 6 (66.67%) | 6 (85.71%) | |
| **MetS components** | | | |
| Obesity | 9 (100.00%) | 5 (71.43%) | 0.175 [b] |
| Dyslipidemia | 9 (100.00%) | 6 (85.71%) | 0.437 [b] |
| Hypertension | 8 (88.89%) | 4 (57.14%) | 0.262 [b] |
| Hyperglycemia | 3 (33.33%) | 2 (28.57%) | 1.000 [b] |
| **Physical activity**, MET-hours/week | 14.39 ± 21.12 | 6.57 ± 6.66 | 0.364 [a] |
| **Smoking** | | | 0.780 [b] |
| Never | 6 (66.67%) | 4 (57.14%) | |
| Former | 1 (11.11%) | 0 (0.00%) | |
| Current | 2 (22.22%) | 3 (42.86%) | |
| **Alcohol consumption** | | | 1.000 [b] |
| Never | 4 (44.44%) | 4 (57.14%) | |
| Monthly | 1 (11.11%) | 0 (0.00%) | |

| Neuropsychological test | Group | | p value |
|---|---|---|---|
| | Adherent (n=9) | Non-adherent (n=7) | |
| **Total sessions attended** | 18.74 ± 10.67 | 2.24 ± 1.14 | 0.001 [a] |
| **Total time training**, hours | 14.06 ± 8.00 | 1.68 ± 0.86 | 0.001 [a] |
| *Paper-and-pencil* | | | |
| **MoCA** | 25.78 ± 2.39 | 25.00 ± 3.27 | 0.590 [a] |
| **Taylor complex figure** | | | |
| Copy | 35.33 ± 1.00 | 35.00 ± 1.53 | 0.606 [a] |
| Immediate recall | 25.78 ± 6.59 | 24.64 ± 5.71 | 0.723 [a] |
| Delayed recall | 23.33 ± 9.85 | 23.43 ± 4.82 | 0.982 [a] |
| **Hopkins Verbal Learning Test** | | | |
| Learning | 23.56 ± 3.91 | 20.86 ± 3.67 | 0.182 [a] |
| Recall | 7.56 ± 2.30 | 8.43 ± 0.98 | 0.365 [a] |
| Recognition | 21.56 ± 1.51 | 21.00 ± 0.82 | 0.396 [a] |
| **Verbal fluency** | | | |
| Phonemic | 11.56 ± 4.56 | 10.71 ± 2.63 | 0.671 [a] |
| Semantic | 16.78 ± 3.23 | 15.43 ± 1.27 | 0.317 [a] |
| **Digit span** | | | |
| Forward | 6.89 ± 1.62 | 7.14 ± 2.19 | 0.793 [a] |
| Backward | 5.33 ± 1.23 | 6.29 ± 2.29 | 0.301 [a] |
| *Computerized* | | | |
| **Continuous Performance Test** | | | |
| Total correct | 98.00 ± 2.45 | 98.00 ± 1.73 | 1.000 [a] |

| | | | |
|---|---|---|---|
| Weekly | 3 (33.33%) | 3 (42.86%) | |
| Almost everyday | 1 (11.11%) | 0 (0.00%) | |
| **Income** | | | 0. 240 [b] |
| Low [1] | 1 (11.11%) | 0 (0.00%) | |
| Average [2] | 0 (0.00%) | 1 (14.29%) | |
| High [3] | 1 (11.11%) | 4 (57.14%) | |
| Very high [4] | 7 (77.78%) | 2 (28.57%) | |
| Mean time (ms) | 0.54 ± 0.07 | 0.49 ± 0.04 | 0.136 [a] |
| **Flanker task** | | | |
| Total correct | 47.88 ± 0.33 | 47.43 ± 0.54 | 0.053 [a] |
| Mean time (ms) | 1.07 ± 0.29 | 0.84 ± 0.21 | 0.098 [a] |
| **Set-Shifting task** | | | |
| Total correct | 102.56 ± 1.59 | 101.86 ± 1.68 | 0.409 [a] |
| Mean time (ms) | 1.05 ± 0.29 | 0.85 ± 0.27 | 0.185 [a] |
| **N-Back task** | 25.78 ± 3.15 | 26.86 ± 0.69 | 0.392 [a] |
| *Functional abilities* | | | |
| **ECog-II** | 1.20 ± 0.18 | 1.13 ± 0.15 | 0.385 [a] |

Abbreviations: MetS, Metabolic Syndrome; MET, metabolic equivalents of energy expenditure; MoCA, Montreal Cognitive Assessment; ms, milliseconds; ECog-II, revised Everyday Cognition scale.

Notes: *Elementary school or junior high school, up to 9 years of education. **High school, up to 12 years of education. ***University degree/MSc/PhD/Postdoc, more than 13 years of education.

[1] up to 900 euros, [2] 901-1400 euros, [3] 1401-2000 euros, [4] >2000 euros.

[a] Comparisons using t-test. [b] Comparisons using Fisher’s exact test. Mean ± standard deviation and frequency (percentage) are presented for continuous and categorical variables, respectively.

**Supplementary Figure S1.** Association between participants' age and total training hours.

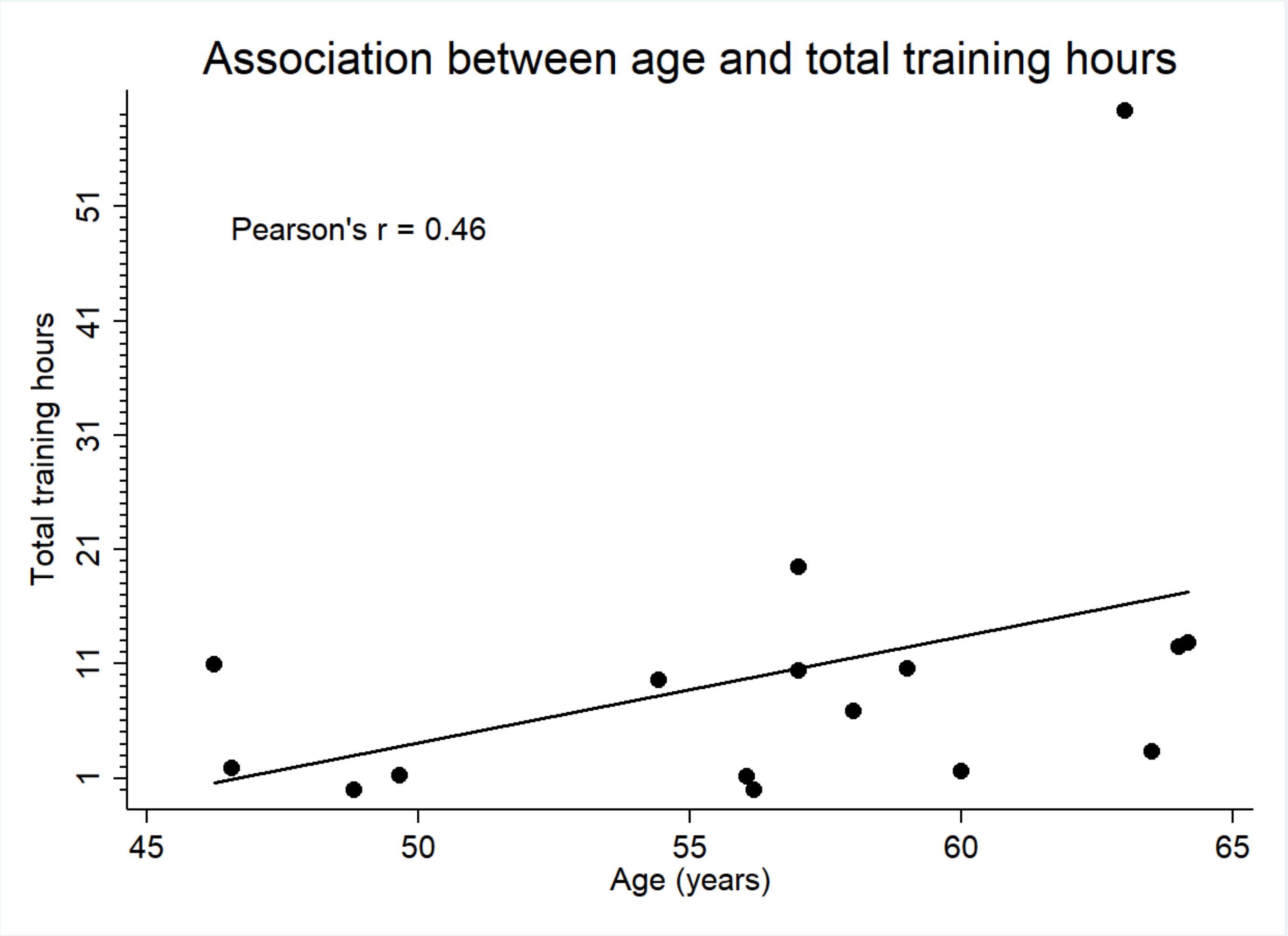

**Supplementary Table S6.** Means and standard deviations for cognitive scores at pre- and post-intervention timepoints of study participants according to intention-to-treat principle.

| Neuropsychological test | Group: Cognitive training (n=16) | Group: Control (n=17) | Neuropsychological test | Group: Cognitive training (n=16) | Group: Control (n=17) |
|---|---|---|---|---|---|
| *Paper-and-pencil* | | | *Computerized* | | |
| **MoCA** | | | **CPT - Total correct** | | |
| Baseline | 25.44 ± 2.73 | 24.06 ± 3.40 | Baseline | 98.00 ± 2.10 | 97.59 ± 2.03 |
| 3-month | 25.31 ± 2.33 | 25.47 ± 2.62 | 3-month | 98.81 ± 1.52 | 98.18 ± 1.47 |
| 1-year | 26.87 ± 1.51 | 24.94 ± 2.19 | 1-year | 98.73 ± 1.10 | 98.65 ± 1.06 |
| **Complex figure - Copy** | | | **CPT - Mean time (ms)** | | |
| Baseline | 35.19 ± 1.22 | 34.35 ± 1.97 | Baseline | 0.52 ± 0.06 | 0.52 ± 0.08 |
| 3-month | 35.63 ± 0.89 | 35.47 ± 0.88 | 3-month | 0.45 ± 0.06 | 0.47 ± 0.08 |
| 1-year | 34.87 ± 2.10 | 35.59 ± 0.80 | 1-year | 0.47 ± 0.07 | 0.50 ± 0.10 |
| **Complex figure - Immediate** | | | **Flanker task - Total correct** | | |
| Baseline | 25.28 ± 6.04 | 25.00 ± 6.81 | Baseline | 47.69 ± 0.48 | 47.47 ± 0.88 |
| 3-month | 28.56 ± 6.10 | 26.71 ± 7.12 | 3-month | 47.75 ± 0.58 | 47.53 ± 0.80 |
| 1-year | 28.33 ± 5.69 | 28.03 ± 6.71 | 1-year | 47.73 ± 0.70 | 47.77 ± 0.44 |
| **Complex figure - Delayed** | | | **Flanker task - Mean time (ms)** | | |
| Baseline | 23.38 ± 7.81 | 23.47 ± 7.98 | Baseline | 0.97 ± 0.28 | 1.02 ± 0.42 |
| 3-month | 27.69 ± 6.12 | 25.62 ± 8.37 | 3-month | 0.85 ± 0.26 | 0.92 ± 0.32 |
| 1-year | 28.20 ± 5.75 | 27.50 ± 7.16 | 1-year | 0.86 ± 0.25 | 0.90 ± 0.23 |
| **HVLT - Learning** | | | **Set-Shifting task - Total correct** | | |
| Baseline | 22.38 ± 3.93 | 22.82 ± 4.83 | Baseline | 102.25 ± 1.61 | 100.06 ± 3.61 |
| 3-month | 23.13 ± 2.50 | 23.35 ± 2.96 | 3-month | 101.25 ± 2.41 | 101.02 ± 3.88 |
| 1-year | 23.13 ± 4.12 | 22.53 ± 4.40 | 1-year | 101.33 ± 1.88 | 100.94 ± 2.36 |

| | | |
|---|---|---|
| **HVLT - Recall** | | |
| Baseline | 7.94 ± 1.84 | 8.35 ± 2.09 |
| 3-month | 8.19 ± 2.20 | 8.88 ± 2.09 |
| 1-year | 7.87 ± 2.48 | 8.00 ± 2.32 |
| **HVLT - Recognition** | | |
| Baseline | 21.31 ± 1.25 | 21.82 ± 1.43 |
| 3-month | 22.25 ± 1.77 | 22.18 ± 1.98 |
| 1-year | 23.13 ± 1.36 | 22.94 ± 1.78 |
| **Verbal fluency - Phonemic** | | |
| Baseline | 11.19 ± 3.75 | 11.41 ± 3.45 |
| 3-month | 12.81 ± 5.52 | 14.00 ± 4.11 |
| 1-year | 11.27 ± 2.05 | 11.29 ± 4.20 |
| **Verbal fluency - Semantic** | | |
| Baseline | 16.19 ± 2.59 | 15.53 ± 3.71 |
| 3-month | 18.63 ± 4.67 | 16.18 ± 4.49 |
| 1-year | 23.60 ± 6.02 | 20.29 ± 6.02 |
| **Digit span - Forward** | | |
| Baseline | 7.00 ± 1.83 | 6.47 ± 2.40 |
| 3-month | 6.88 ± 1.71 | 6.24 ± 2.39 |
| 1-year | 7.07 ± 2.12 | 6.65 ± 2.68 |
| **Digit span - Backward** | | |
| Baseline | 5.75 ± 1.77 | 5.12 ± 2.55 |
| 3-month | 6.38 ± 1.71 | 5.06 ± 2.16 |
| 1-year | 6.60 ± 2.17 | 5.88 ± 2.76 |
| **Set-Shifting task - Mean time (ms)** | | |
| Baseline | 0.96 ± 0.29 | 1.05 ± 0.34 |
| 3-month | 0.83 ± 0.25 | 0.95 ± 0.34 |
| 1-year | 0.89 ± 0.32 | 0.92 ± 0.35 |
| **N-Back task** | | |
| Baseline | 26.25 ± 2.41 | 26.18 ± 2.92 |
| 3-month | 27.44 ± 1.63 | 27.06 ± 1.35 |
| 1-year | 27.00 ± 1.77 | 26.82 ± 2.24 |

Abbreviations: CI, confidence interval; MoCA, Montreal Cognitive Assessment; HVLT, Hopkins Verbal Learning Test; CPT, Continuous Performance Test; ms, milliseconds.

**Supplementary Table S7.** Mixed models results for cognitive scores according to as-treat principle.

| Neuropsychological test | Beta | 95% CI | Effect size, *d* | Neuropsychological test | Beta | 95% CI | Effect size, *d* |
|---|---|---|---|---|---|---|---|
| *Paper-and-pencil* | | | | *Computerized* | | | |
| **MoCA** | | | | **CPT - Total correct** | | | |
| 3-month | -1.22 | -3.15, 0.71 | 0.28 | 3-month | -0.04 | -1.48, 1.40 | 0.08 |
| 1-year | 0.33 | -1.65, 2.30 | 0.61 | 1-year | -0.15 | -1.63, 1.32 | 0.00 |
| **Complex figure - Copy** | | | | **CPT - Mean time (ms)** | | | |
| 3-month | -0.78 | -2.12, 0.57 | 0.34 | 3-month | -0.03 | -0.08, 0.02 | 0.48 |
| 1-year | -0.54 | -1.92, 0.83 | 0.15 | 1-year | -0.02 | -0.07, 0.04 | 0.22 |
| **Complex figure - Immediate** | | | | **Flanker task - Total correct** | | | |
| 3-month | 3.02 | -0.78, 6.82 | 1.03 | 3-month | -0.39 | -1.07, 0.29 | 0.13 |
| 1-year | 1.39 | -2.51, 5.29 | 0.55 | 1-year | -0.44 | -1.14, 0.26 | 0.22 |
| **Complex figure - Delayed** | | | | **Flanker task - Mean time (ms)** | | | |
| 3-month | 3.85 | -0.39, 8.10 | 1.07 | 3-month | -0.07 | -0.23, 0.09 | 0.13 |
| 1-year | 3.02 | -1.34, 7.38 | 0.86 | 1-year | -0.06 | -0.22, 0.11 | 0.07 |
| **HVLT - Learning** | | | | **Set-Shifting task - Total correct** | | | |
| 3-month | -0.57 | -3.86, 2.72 | 0.06 | 3-month | -0.88 | -4.47, 0.72 | 0.16 |
| 1-year | 1.77 | -1.60, 5.14 | 0.73 | 1-year | -1.83 | -5.03, 0.28 | 0.25 |
| **HVLT - Recall** | | | | **Set-Shifting task - Mean time (ms)** | | | |
| 3-month | 0.83 | -0.82, 2.48 | 0.33 | 3-month | -0.07 | -0.25, 0.11 | 0.27 |
| 1-year | 1.66 | -0.03, 3.36 | 0.89 | 1-year | 0.07 | -0.11, 0.26 | 0.64 |
| **HVLT - Recognition** | | | | **N-Back task** | | | |
| 3-month | 0.05 | -1.37, 1.45 | 0.15 | 3-month | 1.49 | -0.55, 3.52 | 0.64 |
| 1-year | 0.93 | -0.52, 2.37 | 0.84 | 1-year | 0.38 | -1.71, 2.47 | 0.05 |
| **Verbal fluency - Phonemic** | | | | | | | |

| | | | |
|---|---|---|---|
| 3-month | 0.44 | -2.51, 3.40 | 0.36 |
| 1-year | 0.30 | -2.73, 3.34 | 0.31 |
| **Verbal fluency - Semantic** | | | |
| 3-month | -0.10 | -4.28, 4.09 | 0.07 |
| 1-year | -0.82 | -5.10, 3.47 | 0.12 |
| **Digit span - Forward** | | | |
| 3-month | 0.10 | -1.15, 1.34 | 0.14 |
| 1-year | 0.10 | -1.18, 1.38 | 0.15 |
| **Digit span - Backward** | | | |
| 3-month | 1.15 | -0.07, 2.37 | 0.90 |
| 1-year | 0.52 | -0.73, 1.77 | 0.33 |

Abbreviations: CI, confidence interval; MoCA, Montreal Cognitive Assessment; HVLT, Hopkins Verbal Learning Test; CPT, Continuous Performance Test; ms, milliseconds.

Notes: Models adjusted for baseline score, age, sex, education and recreational physical activity. p values represent statistical significance of the group x timepoint interaction term.

* p <0.05, **p < 0.003

**Supplementary Figure S2.** Estimated means of paper-and-pencil cognitive scores over time by intervention group according to as-treated principle.

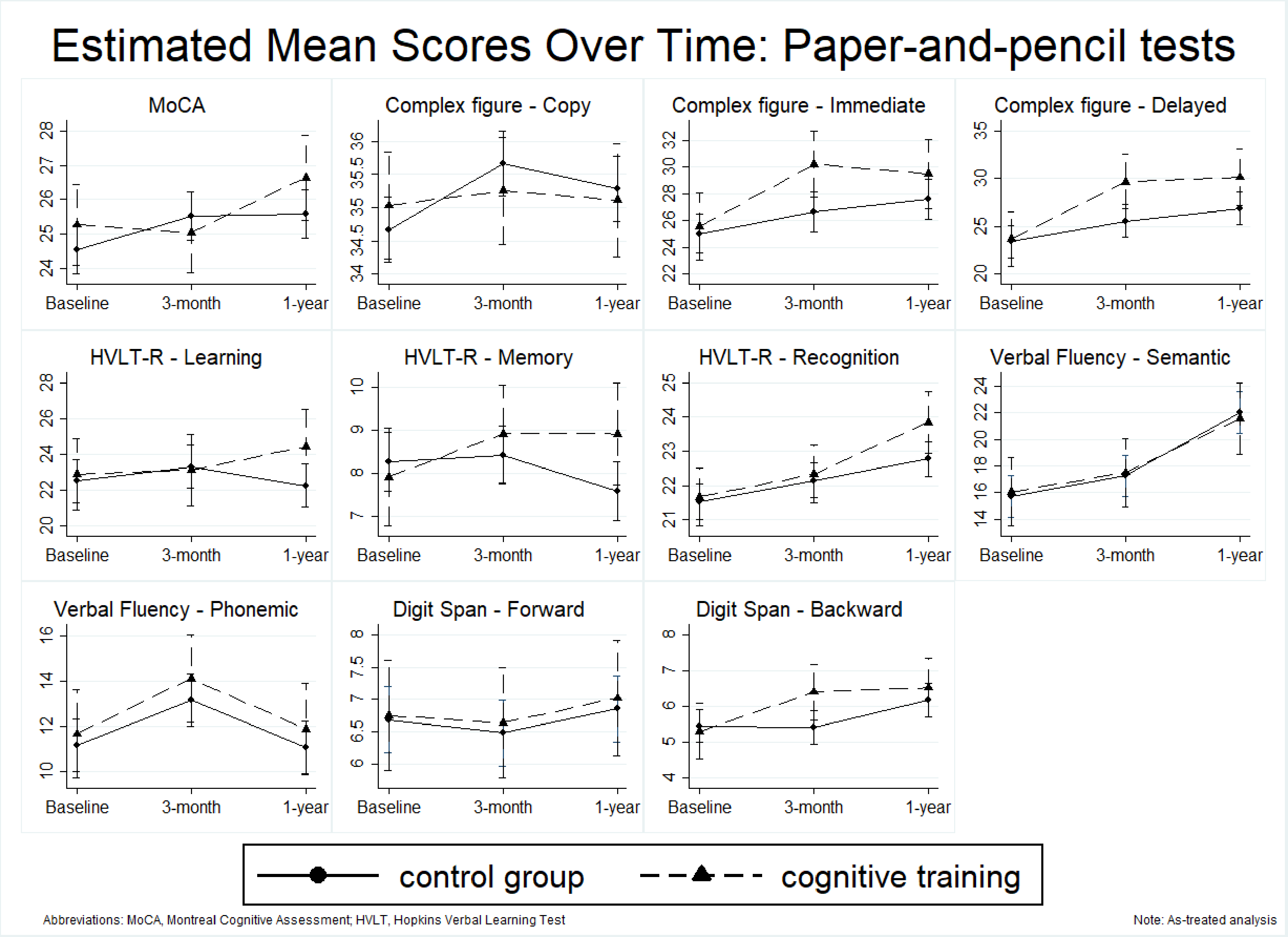

**Supplementary Figure S3.** Estimated means of computerized cognitive scores over time by intervention group according to as-treated principle.

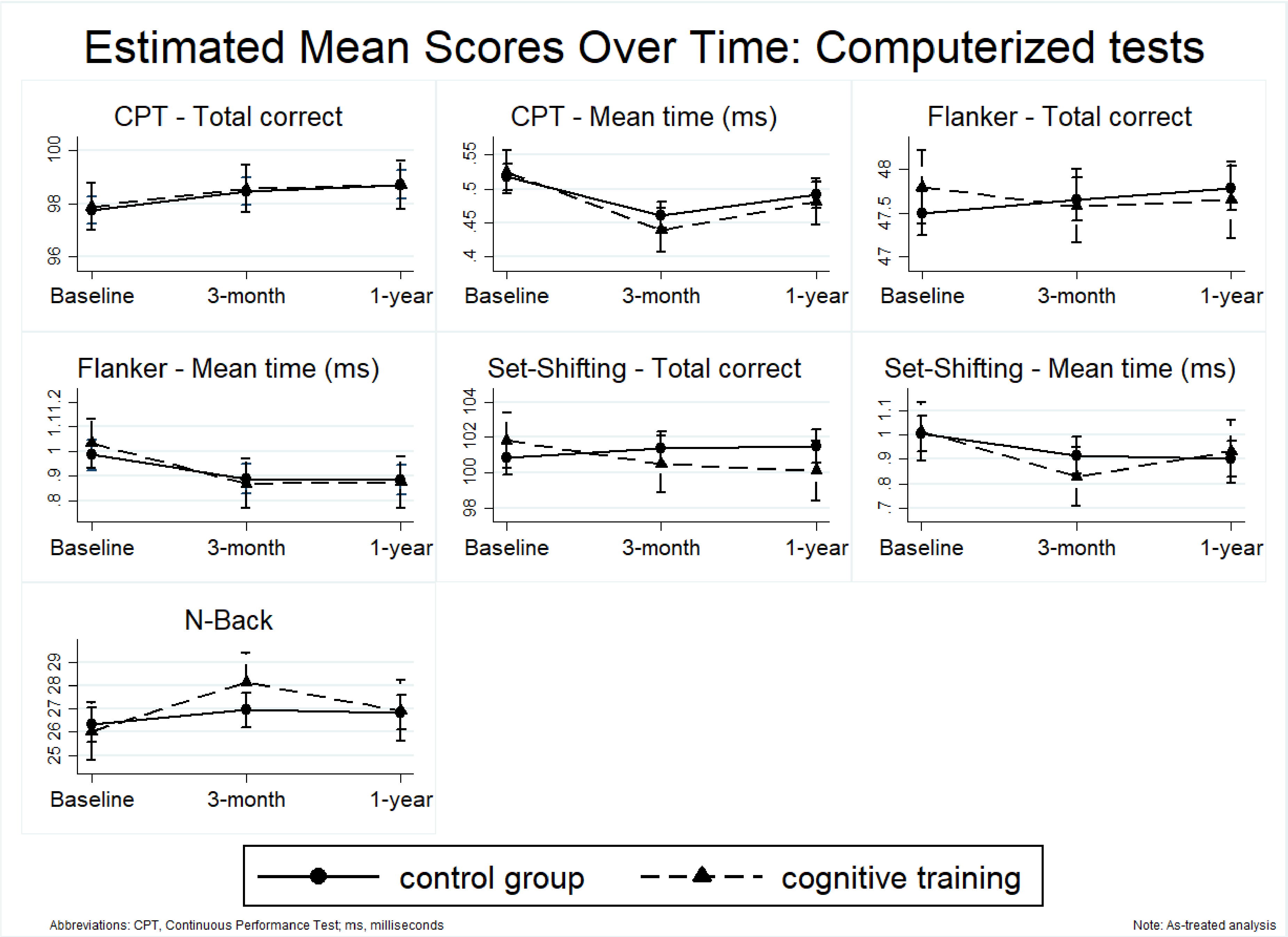

**Supplementary Figure S4.** Associations of participants' age with paper-and-pencil cognitive scores at 3-month follow-up and total training hours.

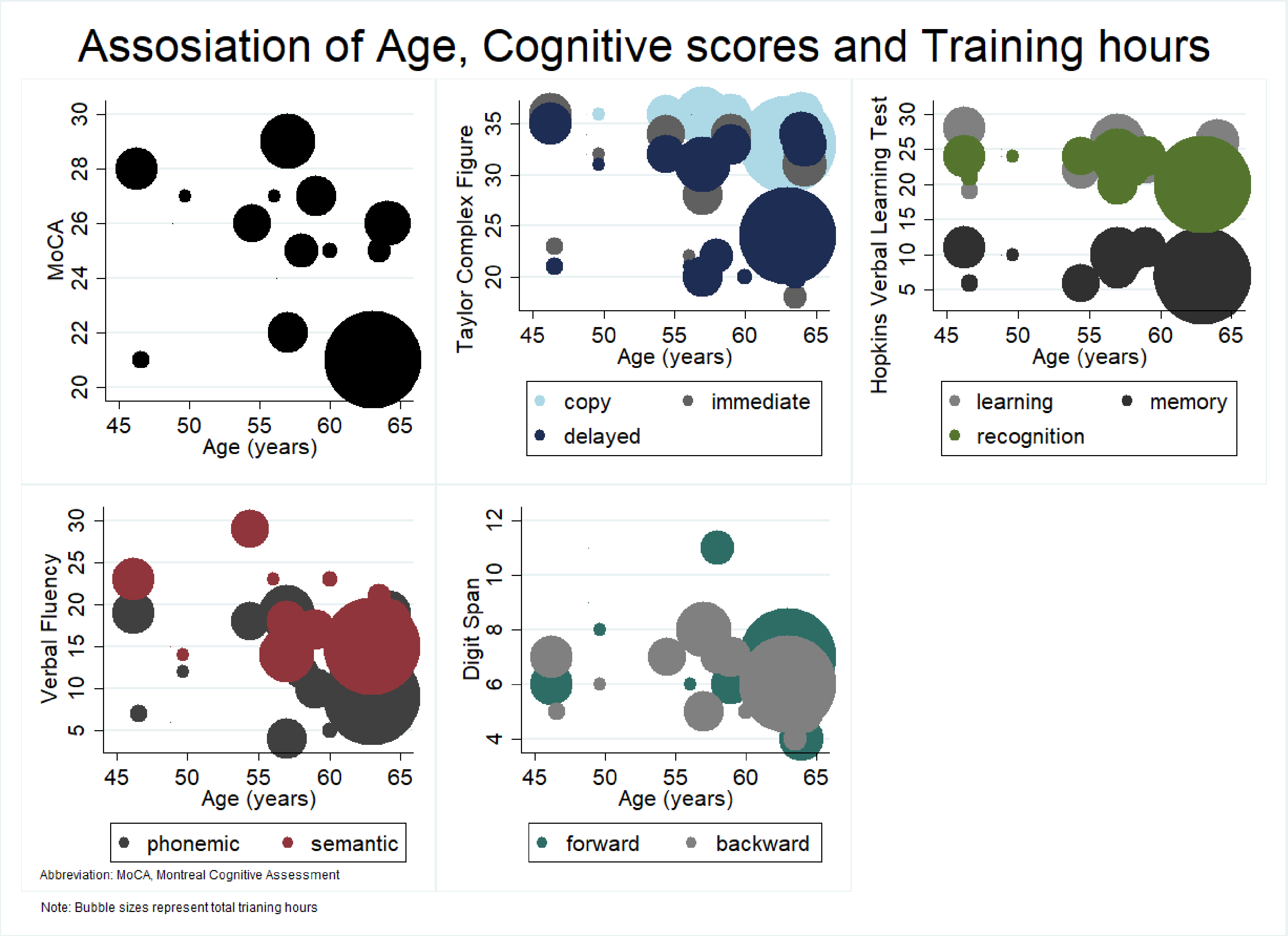

**Supplementary Table S8.** Mean change in cognitive scores at 3-month and 1-year follow-up within groups according to as-treated principle.

| Neuropsychological test | Cognitive training (n=9) | | Control (n=24) | | Neuropsychological test | Cognitive training (n=9) | | Control (n=24) | |
|---|---|---|---|---|---|---|---|---|---|
| | MD (SD) | p value | MD (SD) | p value | | MD (SD) | p value | MD (SD) | p value |
| *Paper-and-pencil* | | | | | *Computerized* | | | | |
| **MoCA** | | | | | **CPT - Total correct** | | | | |
| 3-month | -0.22 (2.68) | 1.000 | 1.00 (3.09) | 0.709 | 3-month | 0.67 (0.71) | 1.000 | 0.71 (2.42) | 0.891 |
| 1-year | 1.25 (2.32) | 1.000 | 1.04 (2.20) | 0.582 | 1-year | 0.88 (2.03) | 1.000 | 0.96 (1.97) | 0.161 |
| **Complex figure - Copy** | | | | | **CPT - Mean time (ms)** | | | | |
| 3-month | 0.22 (1.48) | 1.000 | 1.00 (1.77) | 0.065 | 3-month | -0.09 (0.06) | 0.001 | -0.06 (0.07) | <0.001 |
| 1-year | 0.00 (0.00) | 1.000 | 0.63 (2.28) | 1.000 | 1-year | -0.04 (0.02) | 0.748 | -0.03 (0.06) | 0.694 |
| **Complex figure - Immediate** | | | | | **Flanker task - Total correct** | | | | |
| 3-month | 4.67 (3.16) | 0.059 | 1.65 (4.39) | 1.000 | 3-month | -0.22 (0.83) | 1.000 | 0.17 (1.09) | 1.000 |
| 1-year | 3.88 (4.49) | 0.287 | 2.54 (5.56) | 0.155 | 1-year | -0.13 (0.84) | 1.000 | 0.29 (0.81) | 1.000 |
| **Complex figure - Delayed** | | | | | **Flanker task - Mean time (ms)** | | | | |
| 3-month | 6.00 (6.63) | 0.014 | 2.15 (5.42) | 0.795 | 3-month | -0.16 (0.09) | 0.239 | -0.10 (0.23) | 0.285 |
| 1-year | 6.19 (6.14) | 0.009 | 3.44 (5.46) | 0.029 | 1-year | -0.16 (0.18) | 0.359 | -0.10 (0.25) | 0.217 |
| **HVLT - Learning** | | | | | **Set-Shifting task - Total correct** | | | | |
| 3-month | 0.22 (3.38) | 1.000 | 0.79 (5.11) | 1.000 | 3-month | -1.00 (2.29) | 1.000 | -0.13 (2.47) | 1.000 |
| 1-year | 1.25 (4.77) | 1.000 | -0.25 (3.59) | 1.000 | 1-year | -1.63 (1.92) | 1.000 | 0.17 (2.70) | 1.000 |
| **HVLT - Recall** | | | | | **Set-Shifting task - Mean time (ms)** | | | | |
| 3-month | 1.00 (1.66) | 1.000 | 0.17 (2.12) | 1.000 | 3-month | -0.23 (0.08) | 0.032 | -0.16 (0.25) | 0.008 |
| 1-year | 0.88 (2.36) | 1.000 | -0.67 (2.12) | 1.000 | 1-year | -0.08 (0.14) | 1.000 | -0.16 (0.26) | 0.008 |
| **HVLT - Recognition** | | | | | **N-Back task** | | | | |
| 3-month | 0.67 (1.14) | 1.000 | 0.63 (1.72) | 1.000 | 3-month | 2.11 (3.18) | 0.226 | 0.63 (2.90) | 1.000 |
| 1-year | 2.25 (1.17) | 0.007 | 1.25 (1.68) | 0.010 | 1-year | 0.38 (2.20) | 1.000 | 0.50 (2.90) | 1.000 |

| | | | | |
|---|---|---|---|---|
| **Verbal fluency - Phonemic** | | | | |
| 3-month | 2.45 (4.61) | 0.789 | 2.00 (3.74) | 0.144 |
| 1-year | 0.22 (3.70) | 1.000 | -0.08 (3.30) | 1.000 |
| **Verbal fluency - Semantic** | | | | |
| 3-month | 1.45 (5.59) | 1.000 | 1.54 (4.12) | 1.000 |
| 1-year | 5.50 (5.01) | 0.044 | 6.29 (5.50) | <0.001 |
| **Digit span - Forward** | | | | |
| 3-month | -0.11 (1.05) | 1.000 | -0.21 (1.44) | 1.000 |
| 1-year | 0.25 (1.28) | 1.000 | 0.17 (1.93) | 1.000 |
| **Digit span - Backward** | | | | |
| 3-month | 1.11 (1.27) | 0.491 | -0.04 (1.73) | 1.000 |
| 1-year | 1.25 (2.05) | 0.342 | 0.71 (1.12) | 0.393 |

Abbreviations: MD, mean difference; SD, standard deviation; MoCA, Montreal Cognitive Assessment; HVLT, Hopkins Verbal Learning Test; CPT, Continuous Performance Test; ms, milliseconds.

Notes: Models adjusted for baseline score, age, sex, education and recreational physical activity. p values represent statistical significance derived from Wald test.
